# UPDATE trial Stage 2: a pre-post exploratory analysis of a behavioural support intervention to reduce ultra-processed food intake, increase minimally processed food intake, and increase physical activity in adults living with overweight or obesity

**DOI:** 10.64898/2026.04.01.26349973

**Authors:** Caroline Buck, Samuel J. Dicken, Gabriella N. Heuchan, Rana E. Conway, Adrian C. Brown, Friedrich C. Jassil, Eli Blair, Chaniqua Ranson, Tapiwa Ruwona, Janine Makaronidis, Chris van Tulleken, Claudia A. M. Gandini Wheeler-Kingshott, Rachel L. Batterham, Abigail Fisher

## Abstract

**Introduction:** High consumption of ultra-processed foods (UPF) is associated with adverse health outcomes and weight gain. Despite increasing calls for behavioural strategies to reduce UPF intake, no theory-informed intervention targeting UPF reduction has been evaluated in UK adults in alignment with national dietary guidance. We assessed the feasibility, acceptability, and preliminary behavioural and clinical outcomes of a multi-component intervention designed to reduce UPF consumption (and increase physical activity (PA)/minimally processed food (MPF) intake).

**Methods:** In this exploratory single-arm pre–post study, adults (N=45) living with overweight or obesity and habitual UPF intake ≥50% of total energy were offered a 6-month behavioural intervention following a controlled feeding phase (UPDATE trial, stage 1). The intervention was developed using the Behaviour Change Wheel and Capability, Opportunity, Motivation-Behaviour (COM-B) model and included one-to-one sessions with a behavioural scientist, tailored print and digital materials, peer-support meetings, and a moderated group chat. Feasibility outcomes included uptake, retention, and intervention fidelity. Secondary outcomes included COM-B constructs, dietary intake, PA, clinical and self-reported outcomes, and qualitative feedback.

**Results:** Uptake was 91% (41/45). Retention at 6 months was 68% (28/41), with 83% (34/41) providing follow-up data (intention-to-treat). Median attendance at one-to-one sessions was 86% (interquartile range (IQR): 57-100) with 56% (23/41) attending all sessions (per-protocol). Fidelity to core behaviour change techniques was high.

At 6 months, COM-B scores improved for healthy eating (+7%, standard deviation (SD): 8; p<0.001) and physical activity (+5%, SD: 9; p=0.013). UPF intake decreased by 25% of total energy (95% confidence interval (95%CI): –32, –17), with a corresponding increase in minimally processed foods (+23%; 95%CI: 17, 29). Vigorous physical activity increased (+60 min/week, IQR: 0-180), weekday sitting time decreased (−61 min/day, SD: 110), and weight reduced by 3.8 kg (IQR: –8.5-1.0; p=0.001). Findings were similar in per-protocol analyses. Qualitative data indicated perceived improvements in wellbeing and habit formation.

**Conclusion:** This theory-informed intervention demonstrated good feasibility and acceptability and was associated with improvements in targeted behavioural mechanisms and health-related outcomes. A randomised controlled pilot trial is warranted to evaluate effectiveness and refine implementation.

## Introduction

Globally, poor diet quality has overtaken tobacco smoking as the leading cause of ill health and premature mortality^1^. In the UK, nearly two-thirds of adults live with overweight or obesity^2^, increasing the risk of chronic disease and early death^3^.

A key driver of poor diet and rising rates of obesity has been changes in the food environment^4^, including the rising prevalence and consumption of ultra-processed food (UPF)^5^. Defined by the Nova classification, UPF are created with the purpose of maximising sales and profit, by developing highly palatable and attractive products to displace minimally-processed food (MPF) and outcompete competitor UPF^6^. In the UK, average daily energy intake from UPF consumption is estimated to be nearly 60%^7^, similar to countries such as the United States^7^.

Mounting evidence suggests that diets high in UPF may have unfavourable impacts on energy intake and weight change compared with nutritionally-matched non-UPF diets^8–11^. High UPF consumption is associated with greater risk of a number of non-communicable diseases such as cardiovascular disease, type 2 diabetes and cancer, and premature mortality^12^. Conversely, reducing UPF intake^8–11^ or substituting UPF with MPF^13,14^ has been associated with favourable effects on energy intake, weight, and metabolic health.

Whilst broader changes to the food system are needed to improve access and affordability to healthier foods^15^, individual-level behavioural support may help reduce UPF intake and associated health risks. However, there is a complex interplay of barriers and facilitators that shape eating behaviours including UPF intake, which varies across individuals^16^. Therefore, well-designed, evidence-based behavioural support programmes (BSPs) targeting UPF reduction are required, including detailed evaluation of their potential utility. Studies to date suggest targeted BSPs designed to reduce UPF intake can be effective in reducing UPF intake and/or improving clinical outcomes, including in pregnant females^17,18^, adolescents^19,20^, and adults living with metabolic syndrome^21^, cardiovascular disease^22^, or overweight and obesity^23^. A systematic review of behavioural interventions targeting UPF reduction found that interventions employing techniques such as goal-setting, self-monitoring, feedback, and practical guidance on identifying and replacing UPF can help people to significantly reduce their UPF intake^24^.

To date, no studies have assessed BSPs to reduce UPF intake in UK adults; all interventions have been conducted in North or South America, and mostly in Brazil. Given differences in food cultures and geographical and sociodemographic patterning of UPF intake^7^, BSP requirements may differ across countries. Additionally, evidence to date comes mainly from small or moderate-quality studies, most relying on self-reported outcomes and showing variable completeness in terms of process evaluation and fidelity reporting of the intervention^23,25,26^. The level of detail and specification of behaviour change techniques (BCTs) and theoretical underpinning also varies considerably, limiting understanding of what drives intervention effectiveness and how best to translate these findings to UK populations^27^. Furthermore, no intervention has combined UPF reduction guidance with UK dietary guidance. This is important, because UPF reduction messaging would be integrated into existing healthy eating guidelines, rather than being delivered as a standalone intervention.

Therefore, the aim of this study was to explore the feasibility and acceptability of a targeted BSP designed to support UK adults to reduce UPF intake, increase MPF intake (and increase physical activity (PA) as a secondary outcome) to aid further development of the intervention. The objectives were first, to gather data on feasibility, uptake, retention, and acceptability of the BSP, and second, to gather preliminary data on the impact of the BSP on diet, PA and health outcomes.

## Methods

UPDATE (Investigating the effects of Ultra-Processed versus minimally processed Diets following UK dietAry guidance on healTh outcomEs) was a two-stage study assessing UPF consumption among UK adults living with overweight or obesity. Stage 1 was a 2 x 2 crossover RCT comparing the health effects of MPF and UPF diets following UK healthy dietary guidance. Stage 2 was a 6-month BSP designed to reduce UPF intake and increase PA. The trial protocol describing Stage 1 has been published^28^, with results published in 2025^10^. A separate protocol describing the development and content of the Stage 2 BSP has also been published^29^.

This present study reports an exploratory single-arm pre-post evaluation of Stage 2, comparing outcomes at the end of Stage 2 with the first baseline prior to Stage 1.

Recruitment, inclusion and exclusion criteria are described in Dicken et al.^28^. Briefly, 55 adults with overweight or obesity (body mass index (BMI) ≥25 to <40 kg/m^2^) and habitual UPF intake ≥50% of total energy intake, were recruited from one hospital trust in London, UK. Baseline participant characteristics are reported in Table 1 of Dicken et al.^10^.

**Table 1:**
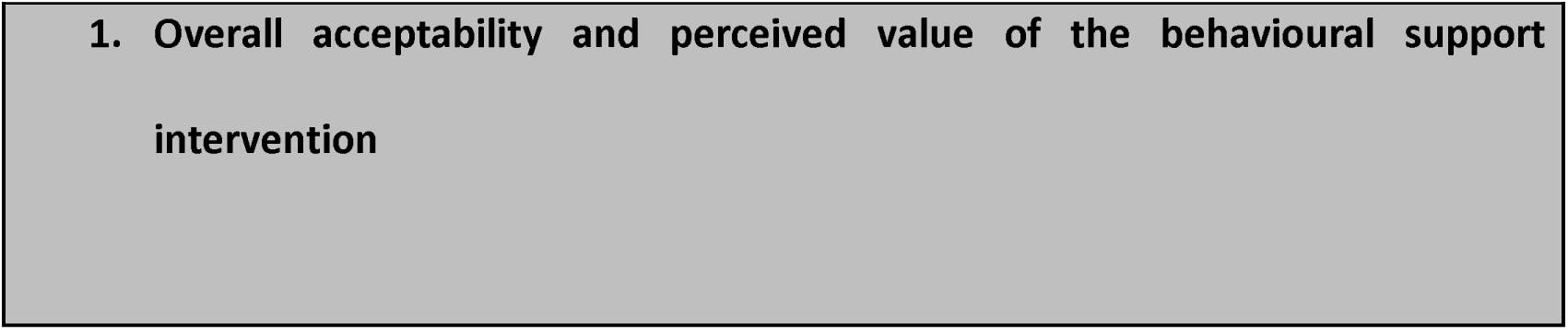
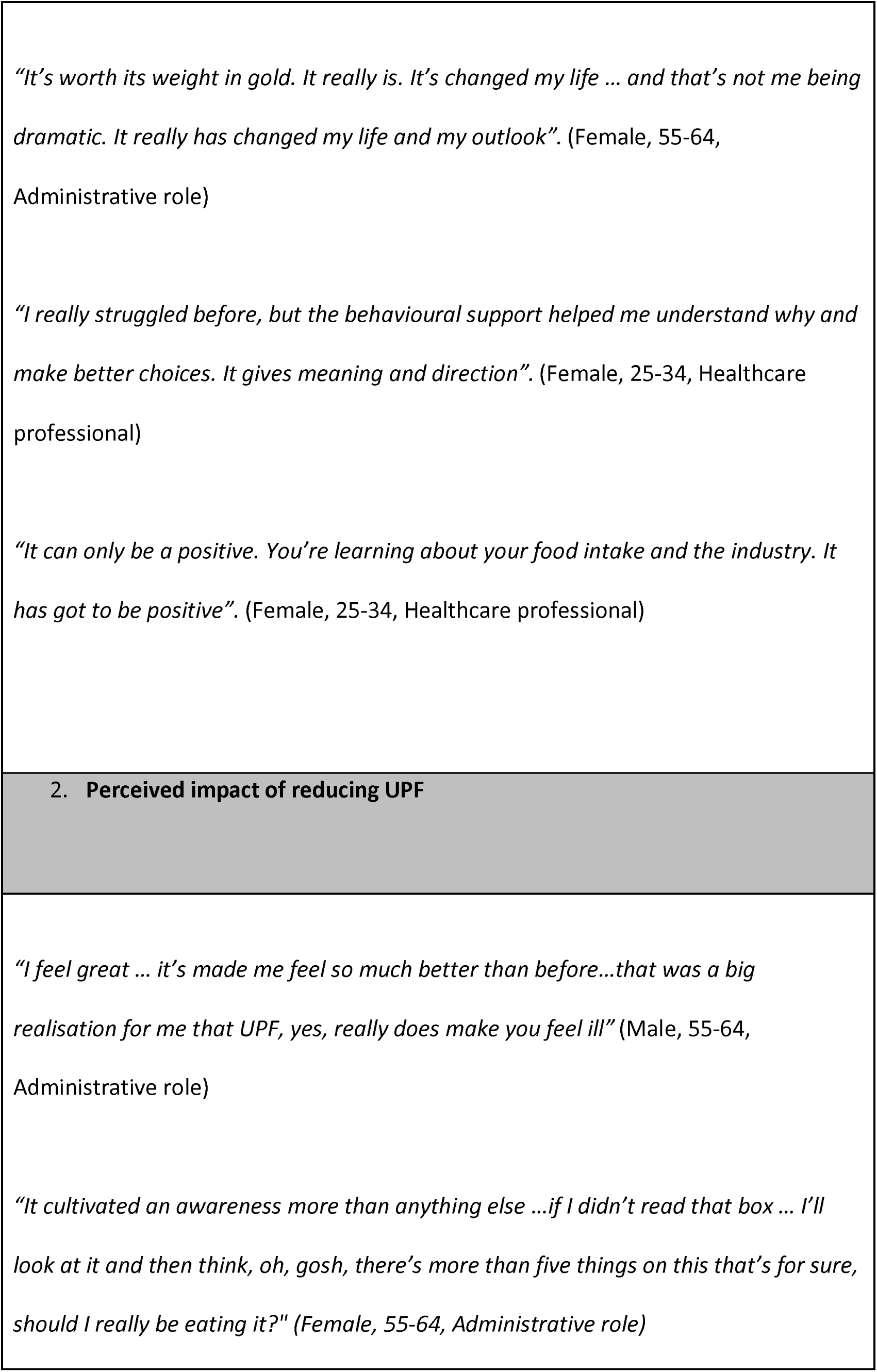
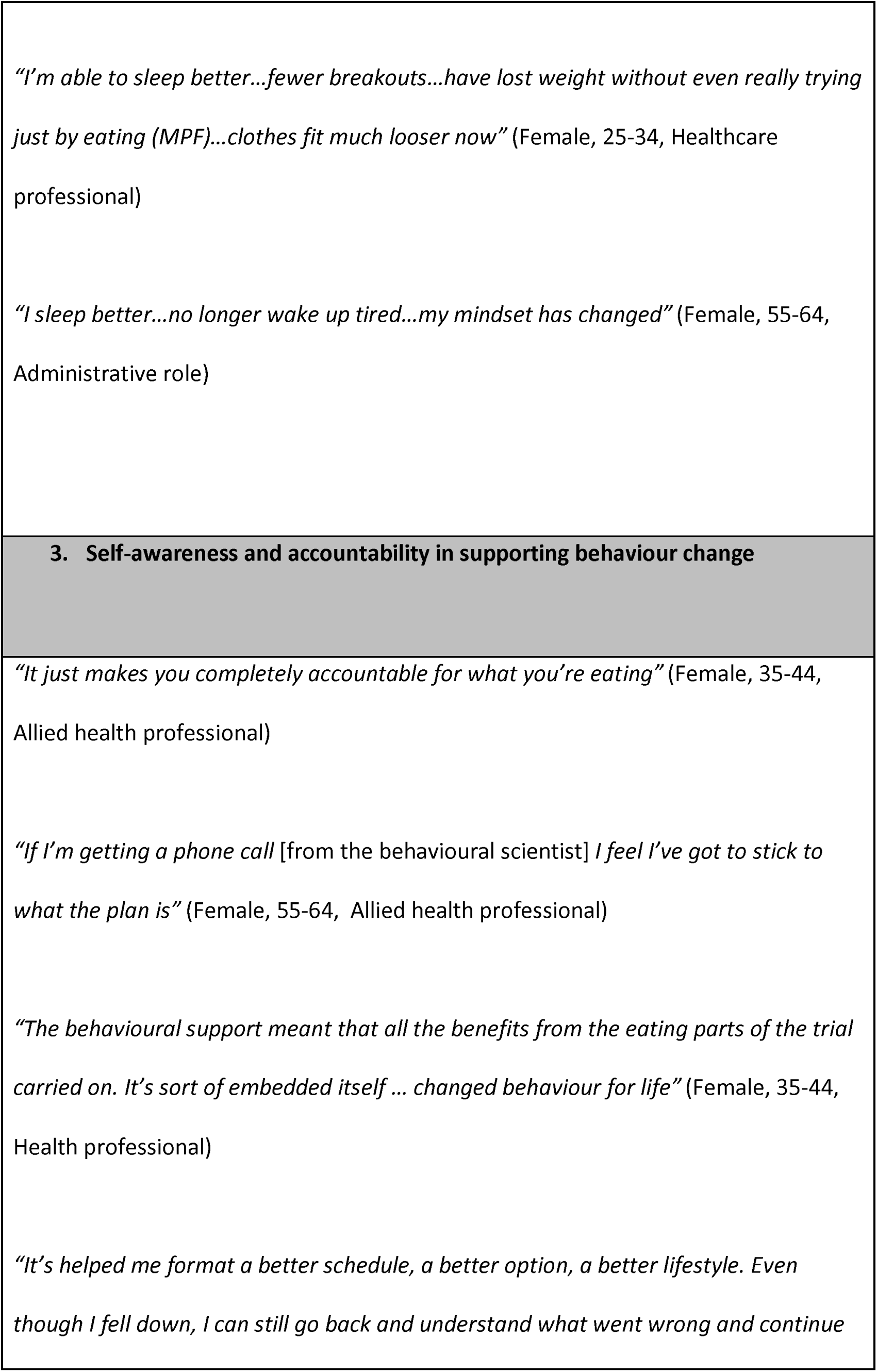
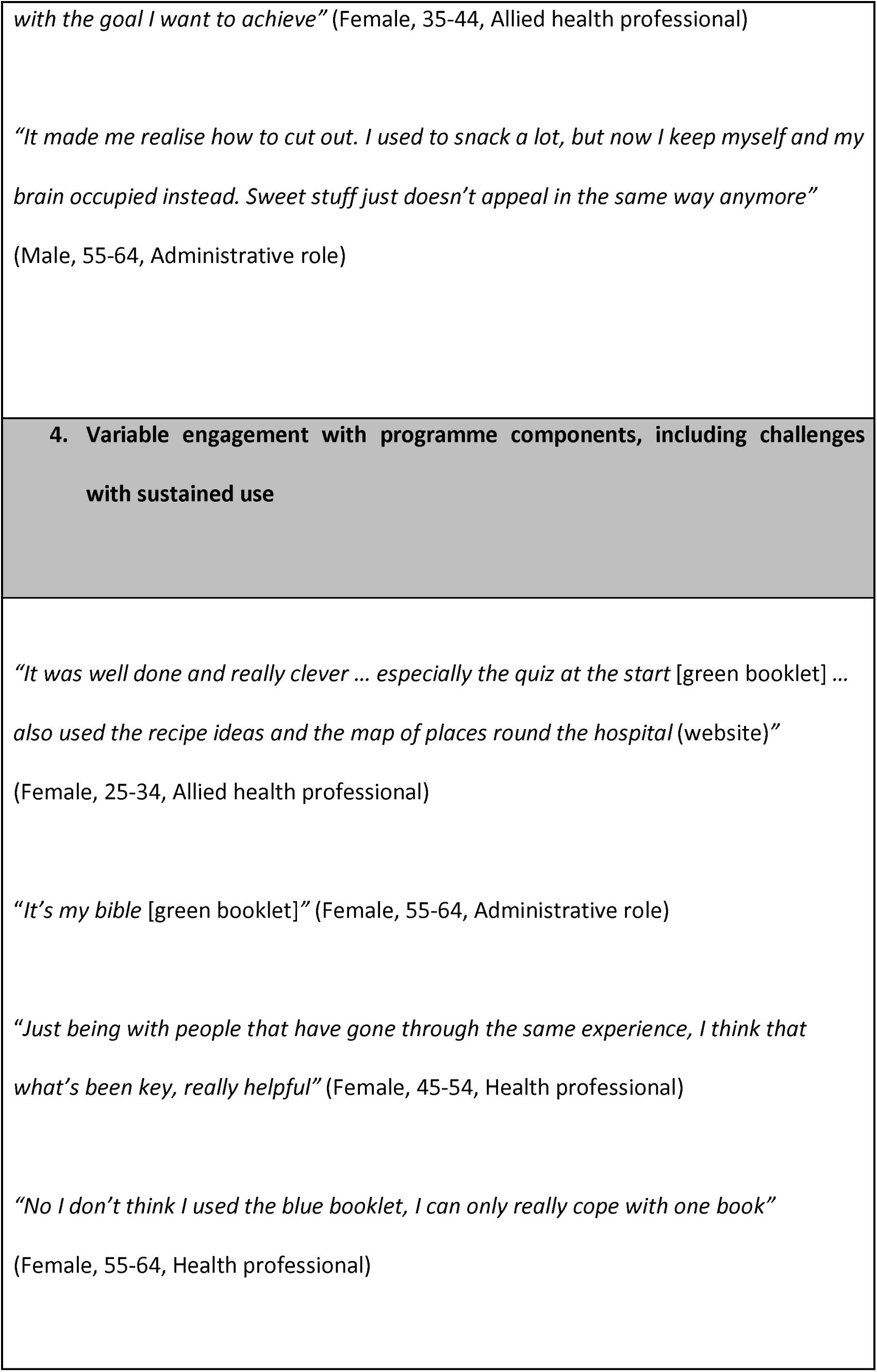
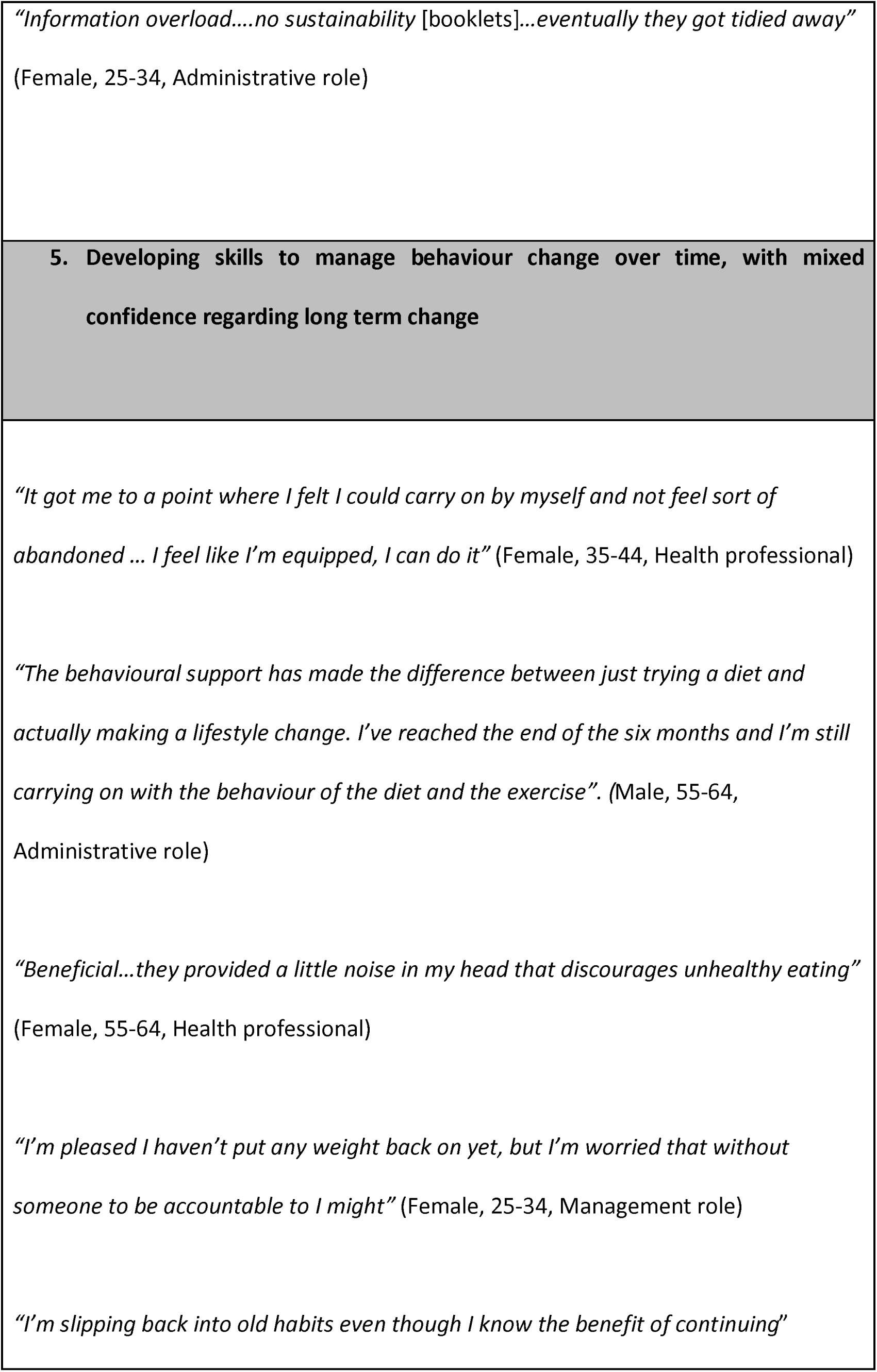
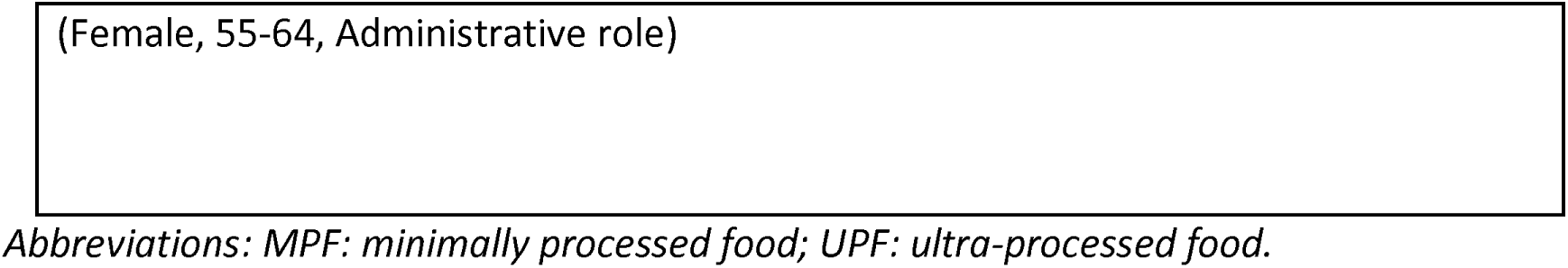
Illustrative quotes of themes capturing participants’ experiences of and responses to the behavioural support programme from exit interviews.

### Intervention

Development and content of the BSP is described in detail in Heuchan et al^29^. Briefly, the intervention was designed to help participants reduce their UPF intake and increase their MPF intake, whilst adhering to the UK Eatwell Guide^30^. The Eatwell Guide provides recommendations on nutrients and foods to consume more of (e.g. fruit, vegetables, fibre) and less of (e.g. foods high in fat, salt and/or sugar). The intervention was designed according to the Medical Research Council (MRC) framework for developing and evaluating complex interventions^31^. The UPDATE BSP was developed using the Behaviour Change Wheel (BCW) framework and the Capability, Opportunity, and Motivation (COM-B) model^32^, linked to the Theoretical Domains Framework (TDF)^33^ to draw on behaviour change theory. A review of existing literature on barriers and facilitators to healthy eating informed identification of relevant behavioural determinants. These barriers and facilitators were mapped onto COM-B components and TDF domains and then linked to relevant intervention functions and BCTs associated with successful dietary change.

### Components

The resulting multi-component programme was delivered over six months and combined online or in-person (by participant choice) one-to-one behavioural support sessions with a behavioural scientist, bespoke print resources for each participant, a website, and regular support groups roughly every six weeks.

Intervention delivery was tailored to individuals. Behavioural scientists reviewed participants’ COM-B questionnaire domains related to healthy eating (collected prior to Stage 1), alongside sociodemographic information, PA, self-reported dietary intake, eating behaviour and mental health measures^29^.

Print resources included two booklets (see Supplemental Material 2 and 3 in Heuchan et al.^29^). The green information booklet provided practical guidance on identifying UPF, the evidence linking UPF with health outcomes, and general dietary advice including the UK Eatwell Guide^30^. The blue tracking booklet was designed to facilitate goal setting and habit formation/tracking.

A mobile-optimised website was created for participants (URL: <u>findmempf.com</u>), which included a recipe bank, a two-week low-UPF meal plan, information on UPF and how to identify them, and the UK Eatwell Guide^30^. Optional online group support sessions and a moderated mobile app group chat were provided to enhance engagement over the 6-month intervention, including a troubleshooting forum, ‘ask the expert’ sessions and opportunities for peer and researcher support.

All sessions were delivered by a team with backgrounds in behavioural science and nutrition, including a health psychologist and researchers in public health and nutrition. All received training in the intervention protocol and behaviour change principles.

### Outcome measures

#### Feasibility

Outcome measures on participant interest, uptake, feasibility, acceptability and retention were collected during and/or at the end of the BSP (6-months follow-up, 49 weeks after recruitment for Stage 1).

Participant interest in the BSP was assessed as the percentage of participants responding yes to ‘*Were you interested in the behavioural support programme when you signed up, or just the (Stage 1) diets*?’ asked at 6-months follow-up.

Uptake was assessed as the percentage of participants agreeing to start the BSP after completing Stage 1. Feasibility was assessed as the percentage of participants receiving the key behavioural support content (defined as the material delivered in the one-to-one behavioural support session in month 1). Retention was assessed as the percentage of participants engaged in the BSP at 6-months follow-up.

#### Acceptability

Acceptability was assessed using several measures. Exit interviews were conducted following the 6-month BSP to explore experiences of participation, acceptability and perceptions. To minimise potential bias, interviews were conducted by team members who had not delivered behavioural support to the participant. Interview transcripts were analysed using qualitative content analysis^34^. Following initial familiarisation, a preliminary codebook was developed inductively by CB based on the analysis of three transcripts. The remaining interviews were analysed by three coders (CB, AF & RC) to capture key aspects of participants’ experiences and views using the agreed codebook, with iterative refinement as analysis progressed. Coded interviews were used to assess the overall acceptability or usefulness of each aspect of the BSP (e.g. one-to-one sessions, website, booklets).

Attendance was defined as the percentage of one-to-one behavioural support sessions a participant was offered that were attended. The withdrawal rate was defined as the percentage of participants who formally withdrew from the BSP.

Lastly, intervention delivery fidelity was assessed to determine whether behavioural support sessions were delivered and implemented as intended, which involved systematic coding of transcribed behavioural support sessions against the session-specific BCT checklists (Supplementary Tables 1-3). The introductory and first two behavioural support sessions were selected for fidelity assessment, as these contained the core intervention content (e.g. education, goal setting, action planning). A purposive sample of seven participants was selected to reflect variation in engagement with the BSP, including participants who completed the programme and those who withdrew, and to ensure representations of sessions delivered by different behavioural scientists. Although seven participants were purposively sampled, one did not receive the month 2 behavioural support session, therefore fidelity coding for that session was based on six transcripts. Transcripts were coded by one behavioural scientist (who had not delivered the intervention) using a structured fidelity framework, with each prespecified BCT rated using a 6-point graded scale adapted from the Dreyfus model of skill acquisition^35^ (Supplementary Table 4), as previously implemented by Cross et al.^36^, allowing assessment of both the presence and quality of BCT delivery. The adapted Dreyfus scale ranged from 0 (poor/absent delivery) to 5 (exceptional delivery), with scores of 3 and 4 reflecting competent and proficient delivery respectively. Four transcripts were double coded by another researcher to ensure consistency.

#### Clinical/patient outcomes

Age, sex, ethnicity, occupation, work pattern, educational level, and marital status were self-reported at screening. Clinical and patient outcomes (weight, waist circumference (WC), BMI, body composition, heart rate, blood pressure, biomarkers, eating behaviour, mental health and quality of life, sleep, and physical fitness) were collected at baseline (week 1-2) and at 6-months follow-up (week 49), and are described in detail previously^10,28^ and in the Supplementary Materials.

Diet (Nova classification groups, total energy, nutrient/food group intake) and alcohol intake were assessed using 24-hour recall (Intake24)^37^. Four non-consecutive recalls were completed at baseline, and two non-consecutive recalls at 6-months follow-up. PA was assessed using the International Physical Activity Questionnaire Short Form (IPAQ-SF)^38^.

#### Analysis

The BSP was not designed to be statistically powered to detect significant changes in outcomes from Stage 1 baseline to 6-months follow-up. Therefore, changes in outcomes are considered exploratory.

Analysis was first conducted on an intention-to-treat (ITT) basis. The ITT sample included all participants providing follow-up data at the end of Stage 2. Analyses were then repeated for participants with 100% attendance at one-to-one sessions (per-protocol (PP)).

Parametrically distributed variables were described using means and standard deviations (SD). Non-parametrically distributed variables were described using medians and interquartile ranges (IQR). Categorical variables were described using counts and percentages.

Changes in aspects of behaviour (COM-B questionnaire), diet, PA, clinical and patient outcomes between baseline and 6-months follow-up were analysed using paired t-tests (parametric) or Wilcoxon tests (non-parametric) where appropriate.

Statistical significance was set at p < 0.05. Analyses were conducted in R (V.2024.12.1+563).

## Results

### Interest, Uptake, Retention

Uptake was 91%; 45 participants were contacted to participate in the BSP after Stage 1, and 41 participants (91%) commenced the BSP. Baseline characteristics of participants commencing the BSP are reported in Supplementary Table 5. Average age at screening was 43.2 years (SD: 11.4), 30 (73.2%) participants were of White ethnicity, 38 (93.7%) were female, and five (12.2%) were night-shift workers. Average baseline weight and BMI were 90.4 kg (SD: 12.8) and 32.6 kg/m^2^ (SD: 4.0).

Retention was 68%; of the 41 participants commencing the BSP, 13 participants formally withdrew or stopped responding (Figure 1). Of these, nine participants formally withdrew (with four providing follow-up data), and four participants dropped out (stopped responding to the behavioural scientist; with two providing follow-up data).

**Figure 1:**
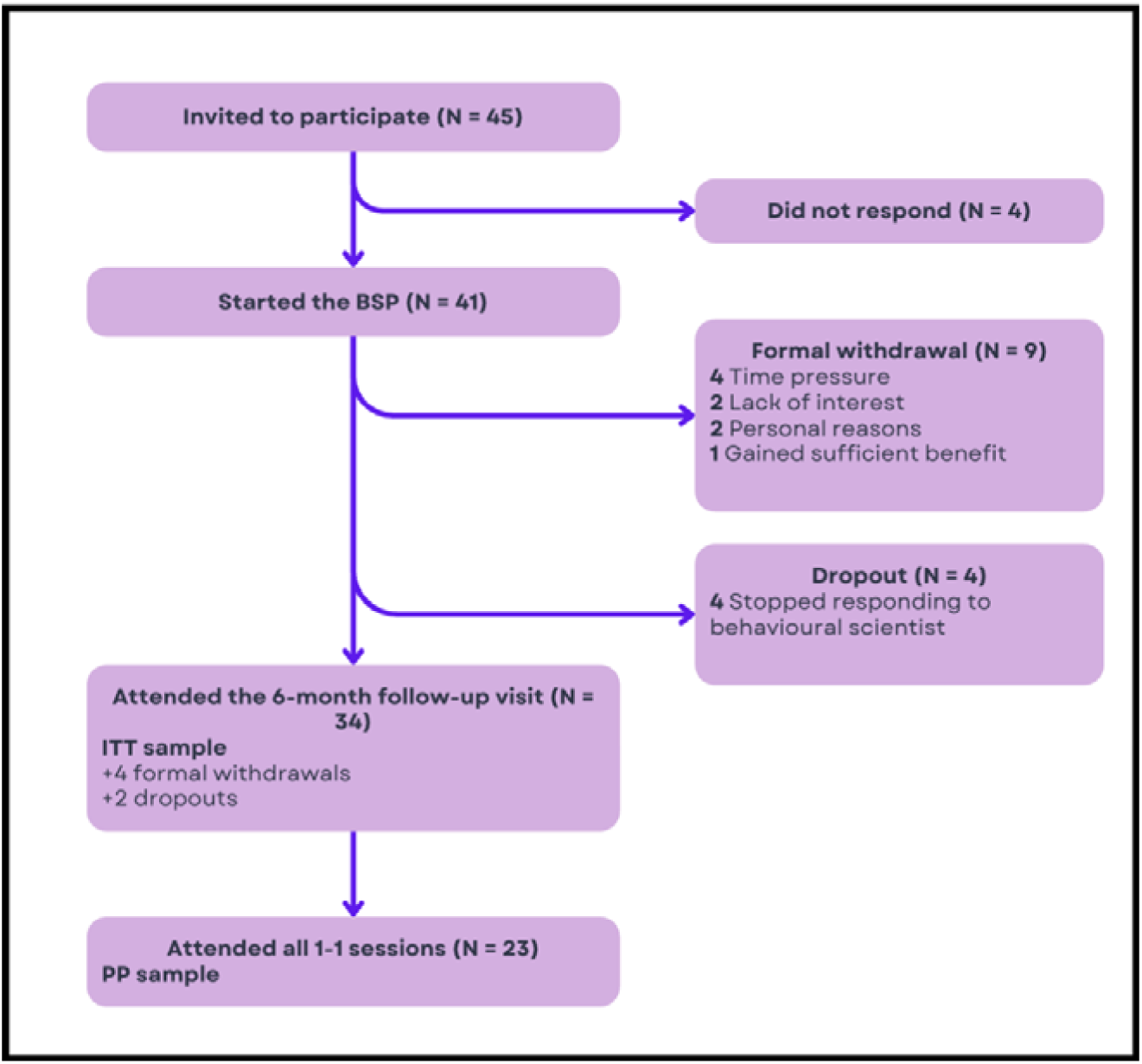
CONSORT diagram of participant flow through the behavioural support programme. ITT = 23 + 4 withdrawals providing follow-up data, +2 dropouts providing follow-up data, +5 participants completing fewer than 100% of one-to-one sessions. PP = no dropouts/withdrawal and attending 100% of one-to-one sessions. Abbreviations: ITT: intention to treat, PP: per protocol, BSP: behavioural support programme.

34 (83%) participants attended the 6-month follow-up visit (ITT sample), of which 31 also completed an exit interview. Median attendance at one-to-one sessions in the ITT sample was 86% (IQR: 57-100). In addition to withdrawals/dropouts, five participants in the ITT sample missed one or more one-to-one sessions, resulting in 23 (56%) participants attending 100% of one-to-one sessions (PP sample).

Baseline characteristics of ITT and PP samples are reported in Supplementary Table 6 and 7, respectively, and of dropouts and withdrawals in Supplementary Table 8. Baseline characteristics between dropouts/withdrawals and ITT were similar.

Of the 31 ITT participants who completed an exit interview, 13 (41.9%) reported they had been interested only in Stage 1 at sign-up, and 17 (54.8%) reported interest in both Stage 1 and the BSP. One participant (3.2%) reported they had been interested in the BSP only and had not realised the study included a dietary phase.

### Feasibility and acceptability

Feasibility was 91% (41/45), defined as receiving the key behavioural support content.

The fidelity assessment indicated that at the session level, overall delivery fidelity was high, particularly in the introductory (mean: 4.4, SD: 1.5) and month 1 sessions (mean: 4.1, SD: 1.3) (Supplementary Table 9 and 10). Fidelity declined in month 2 sessions to within the competent range (mean: 3.4, SD: 1.4), with greater variability across BCTs (Supplementary Table 11).

At the individual BCT level, several techniques demonstrated consistently high delivery fidelity when delivered earlier in the intervention. In the introductory calls, ‘credible source’, ‘review behaviour goals’, ‘behavioural practice’, ‘habit formation’, and ‘social support (emotional and practical)’ were delivered with an ‘expert’ level of fidelity (mean score of 5.0; Supplementary Table 9). In the month 1 sessions, high fidelity was observed for ‘adding objects to the environment’ (5.0), ‘instruction on how to perform the behaviour’ (4.9), ‘self-monitoring of behaviour’ (4.9), and ‘goal setting (behaviour)’ (4.9) (Supplementary Table 10).

In contrast, lower fidelity was observed for some BCTs delivered later in the intervention. In the month 2 sessions, ‘habit formation’ (1.3) and ‘feedback on outcome of behaviours’ (1.7) demonstrated the lowest fidelity scores (Supplementary Table 11). Inter-rater reliability was strong, with 90% agreement across double-coded transcripts.

### Acceptability

Analysis of exit interviews identified key themes capturing participants’ experiences of and responses to the BSP. Representative quotes are provided in Table 1.

#### 1. Overall acceptability and perceived value

Participants consistently described the BSP as acceptable and worthwhile overall. Many reported that it felt supportive rather than prescriptive, and the length and pacing of the programme was largely manageable alongside daily commitments. Acceptability was commonly framed in pragmatic terms, with participants recognising that not all components suited them, but that the intervention was worth the time and effort required.

#### 2. Perceived impact of reducing UPF

Participants frequently linked reducing UPF intake with improvements in physical and emotional wellbeing. These included feeling better day-to-day, having increased energy, experiencing more clarity, and noticing mood improvements. Several participants reported feeling more in control of their eating, which appeared to reinforce motivation to continue reducing UPF. Participants varied in how they described these changes; some spoke about specific physical or emotional effects, while others described broader shifts in their thoughts about food and its role in their health. Some also reported changes in how they perceived themselves in relation to food, including adopting healthier eating patterns as part of their sense of identity. Participants also described developing a clearer understanding of UPF and learning how to identify and replace UPF in ways that felt manageable long term.

#### 3. Self-awareness and accountability in supporting behaviour change

Participants commonly described increased awareness of eating habits and triggers for eating UPF and other dietary behaviours. Many linked this awareness to accountability created through the BSP, particularly the expectation of reflecting on choices and discussing progress and difficulties during the sessions with the behavioural scientist. Accountability was not described as pressurising or negative, but as supportive, helping participants to stay engaged, acknowledge progress and resetting following lapses. One-to-one sessions with a behavioural scientist were frequently cited as a valued part of the intervention and appeared central to these processes. Participants highlighted the tailored and non-judgmental nature of the support, alongside the opportunity to problem-solve and adapt strategies to their own circumstances.

#### 4. Variable engagement with programme components, including challenges with sustained use

Engagement with specific components varied. While some found the printed materials, website, and peer group meetings useful, others reported minimal or short-term use. The blue tracking booklet was often described as burdensome, with the vast majority reporting stopping using it after the introductory session. The website was more consistently identified as useful, particularly for recipes and the ‘food finding’ component. Engagement with group sessions and expert input also differed, often depending on preference and availability.

#### 5. Developing skills to manage behaviour change over time, with mixed confidence regarding long-term change

Many participants reported developing skills and ways of thinking that they felt could support dietary change beyond reducing UPF intake, and for some, increased PA. This included coping with lapses, planning ahead and applying goal setting approaches incrementally. Whilst several described feeling optimistic that learned approaches to behaviour change would be sustainable after the BSP, others were less certain about maintaining changes without continued support.

### COM-B

Baseline COM-B healthy eating questionnaire scores and changes in COM-B scores are reported in Table 2 (ITT) and (Supplementary Table 12 (PP)). At 6-months follow-up, there was a significant improvement in COM-B healthy eating total score (7.3%, SD: 8.4), p < 0.001), knowledge, action control, action planning, habit, environment, identity, motivation and reinforcement from baseline. Results were consistent in the PP sample (Supplementary Table 12).

**Table 2:**
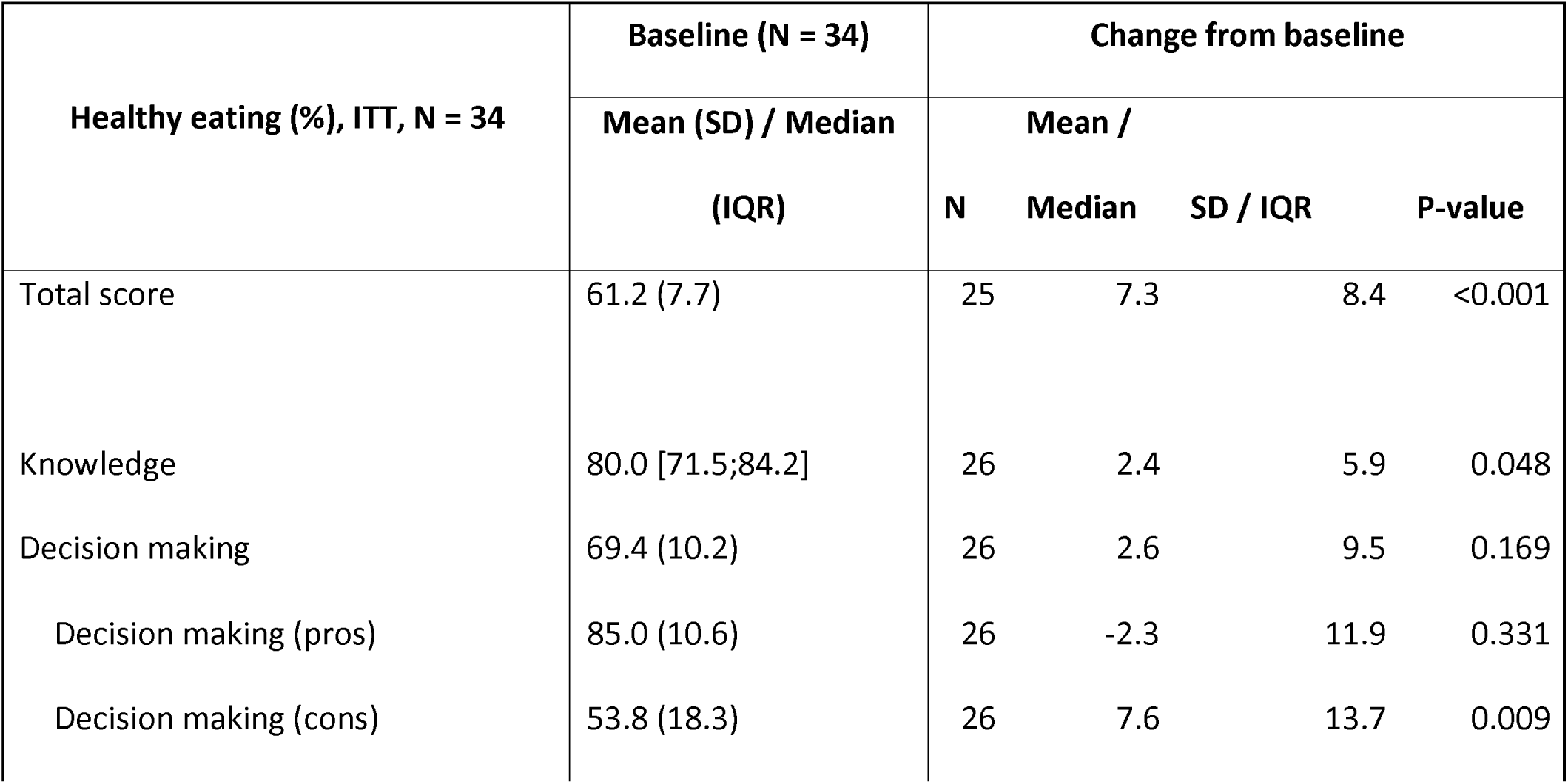
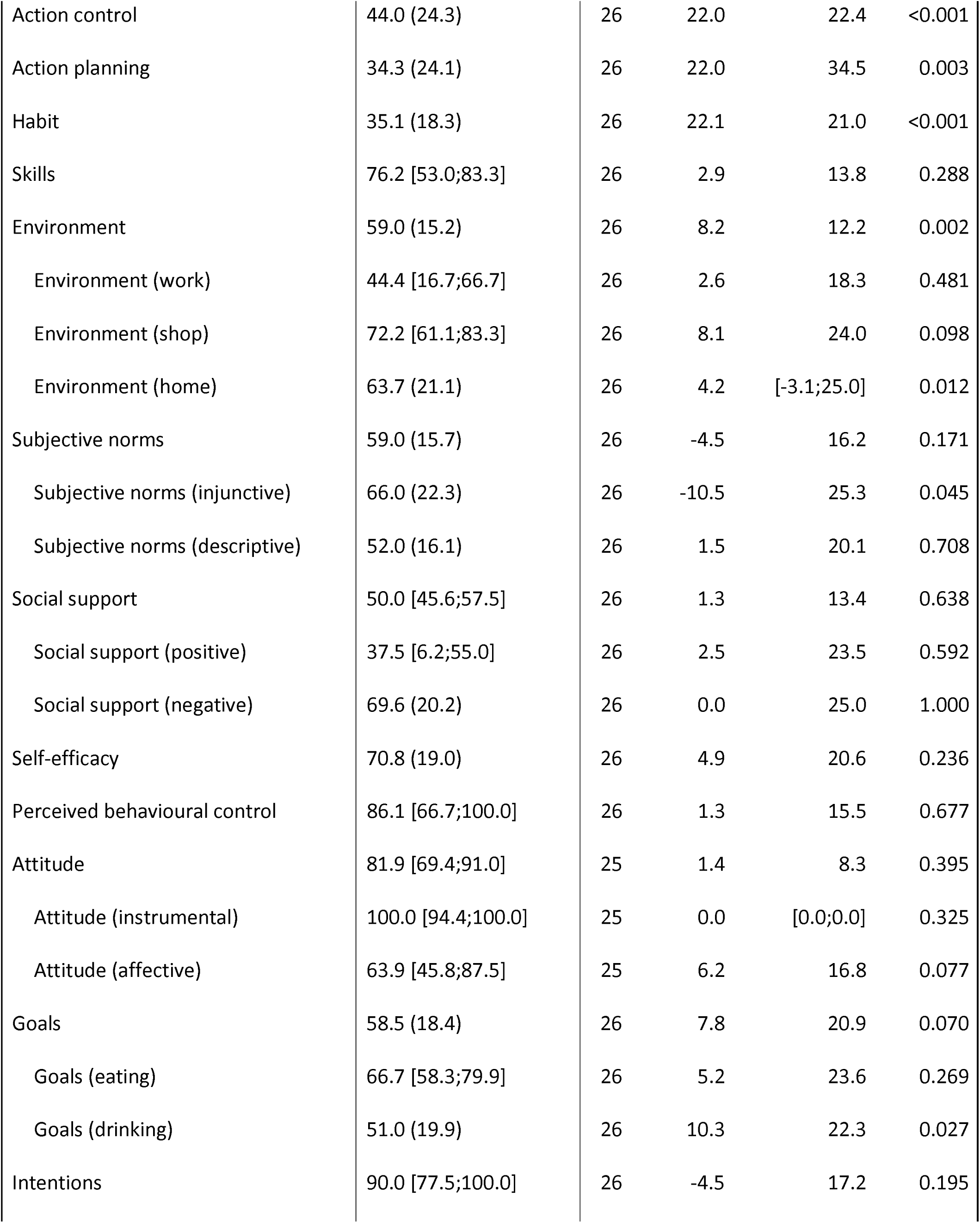
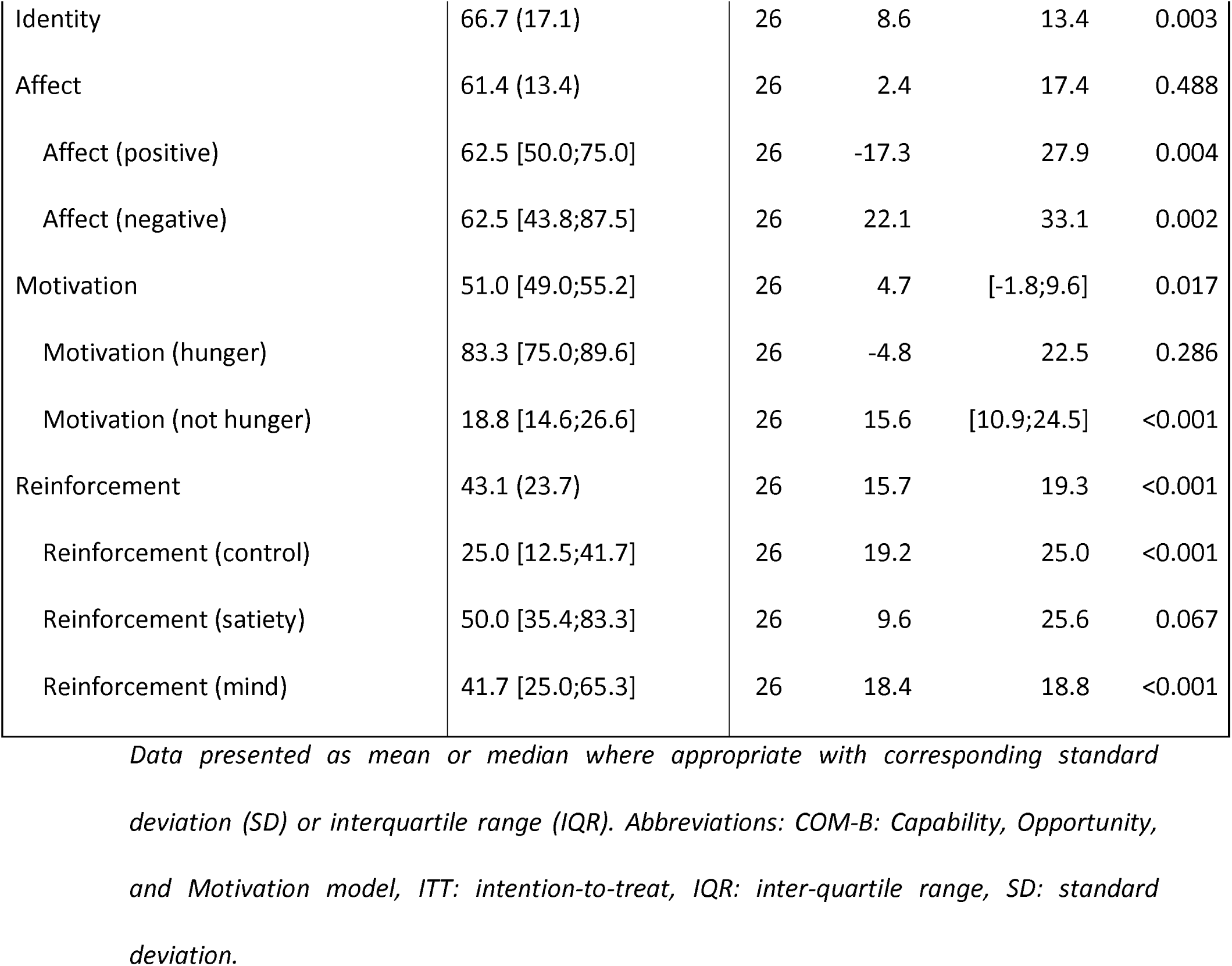
ITT: COM-B healthy eating questionnaire scores at baseline, and changes in COM-B healthy eating scores from baseline to 6-month follow-up.

Baseline COM-B PA questionnaire scores and changes in COM-B PA scores are reported in Supplementary Tables 13 (ITT) and 14 (PP). At 6-months follow-up, there was a significant improvement in COM-B PA total score (+4.8%, SD: 9.0, p = 0.013), action control, habit, skills, affect and reinforcement from baseline. At 6-months follow-up, there was a significant reduction in goals from baseline. Results were consistent in the PP sample (Supplementary Table 14).

### Diet

Changes in self-reported dietary intake are reported in Supplementary Table 15 (ITT) and 16 (PP). From baseline, there was a significant decrease in total energy (−823 kcal (SD: 608), p < 0.001), UPF (−911 kcal (SD: 622); –24.8 % kcal (SD: 20.0), both p < 0.001), processed culinary ingredients (PCI) (−27 kcal (SD: 67), saturated fat (−1.9 % kcal (SD: 4.5), p = 0.035), carbohydrate (−2.9 % kcal (SD: 6.1), p = 0.016), free sugar (−3.6 % kcal (SD: 4.5), p < 0.001), salt (−3.0 g (SD: 2.4), p < 0.001), alcohol (0.0 % kcal (IQR: –1.7; 0.0), p = 0.025) and fibre (−5.4 g (SD: 5.8), p < 0.001) intake at 6-months follow-up (Table 3). MPF (149 kcal (SD: 234), p = 0.002; 22.6 % kcal (SD: 16.0), p < 0.001), and protein (5.1 % kcal (SD: 4.3), p < 0.001) intake significantly increased at 6-months follow-up. Results were consistent in the PP sample.

**Table 3:** ITT: Changes in clinical and patient-reported outcomes from baseline to 6-month follow-up.

| Clinical and patient-reported outcomes, ITT,<br>N = 34 | Change from baseline |  |  |  |
| --- | --- | --- | --- | --- |
|  | N | Mean /<br>Median | SD / IQR | p-value |
| Percentage weight change (%) | 34 | -3.5 | [-8.6;1.1] | <0.001 |
| Weight (kg) | 34 | -3.0 | [-8.5;1.0] | 0.001 |
| Body mass index (kg/m<sup>2</sup>) | 34 | -1.0 | [-2.8;0.3] | 0.001 |
| Waist circumference (cm) | 34 | -0.5 | [-4.8;2.0] | 0.294 |
| Waist-to-height ratio | 34 | -0.003 | [-0.03;0.01] | 0.243 |
| Fat mass (kg) | 34 | -2.3 | [-5.6;0.9] | <0.001 |
| Body fat percentage (%) | 34 | -1.5 | 2.3 | 0.001 |
| Visceral fat rating | 34 | -1.0 | [-1.0;0.0] | 0.001 |
| Fat-free mass (kg) | 34 | -0.8 | [-2.6;0.2] | 0.009 |
| Muscle mass (kg) | 34 | -0.8 | [-2.4;0.2] | 0.010 |
| Bone mass (kg) | 34 | 0.0 | [-0.1;0.0] | 0.007 |
| Total body water (kg) | 34 | -1.0 | [-2.1;0.0] | <0.001 |
| Total body water percentage (%) | 34 | 0.9 | 1.9 | 0.006 |
| Estimated BMR | 34 | -31.0 | [-92.5;11.8] | 0.003 |
| Systolic blood pressure (mmHg) | 33 | -8.1 | 13.2 | 0.001 |
| Diastolic blood pressure (mmHg) | 33 | 0.0 | [-7.0;2.5] | 0.368 |
| Resting heart rate (bpm) | 33 | -5.3 | 10.1 | 0.005 |
| Bilirubin (umol/L) | 34 | 0.4 | 2.9 | 0.483 |
| Alkaline phosphatase (IU/L) | 29 | 0.4 | 7.8 | 0.760 |
| Alanine transaminase (IU/L) | 26 | -2.0 | [-4.8;0.8] | 0.022 |
| Albumin (g/L) | 34 | 0.8 | 1.8 | 0.013 |
| HbA1C (%) | 33 | -0.1 | 0.3 | 0.113 |
| Fasting glucose (mmol/L) | 33 | -0.1 | [-0.3;0.2] | 0.209 |
| Total cholesterol (mmol/L) | 33 | 0.1 | 0.6 | 0.613 |
| Total cholesterol:HDL ratio | 33 | -0.1 | 0.5 | 0.151 |
| HDL-C (mmol/L) | 33 | 0.0 | [-0.1;0.2] | 0.308 |
| LDL-C (mmol/L) | 33 | 0.1 | 0.5 | 0.492 |
| Non-HDL-C (mmol/L) | 33 | 0.0 | 0.6 | 1.000 |
| Triglycerides (mmol/L) | 33 | -0.1 | [-0.4;0.1] | 0.126 |
| C-reactive protein (mg/L) | 32 | -0.3 | [-1.2;0.1] | 0.017 |
| PoF - Food available | 27 | -0.8 | [-1.4;-0.7] | <0.001 |
| PoF - Food present | 27 | -1.2 | 1.1 | <0.001 |
| PoF - Food tasted | 27 | -0.6 | [-1.0;-0.2] | <0.001 |
| PoF – Total | 27 | -0.9 | [-1.3;-0.4] | <0.001 |
| CoEQ - Craving control | 27 | 25.0 | 20.1 | <0.001 |
| CoEQ - Craving for sweet | 27 | -12.9 | 10.7 | <0.001 |
| CoEQ - Craving for savoury | 27 | -18.4 | 17.8 | <0.001 |
| CoEQ - Positive Mood | 27 | 8.7 | 20.6 | 0.037 |
| CoEQ - How difficult has it been to resist eating this food during the last 7 days? | 27 | -27.5 | 27.7 | <0.001 |
| IWQoL Lite - Physical function | 29 | 14.1 | 15.1 | <0.001 |
| IWQoL Lite - Self esteem | 28 | 23.5 | 20.0 | <0.001 |
| IWQoL Lite - Sexual life | 28 | 11.6 | 21.8 | 0.009 |
| IWQoL Lite - Public distress | 29 | 0.0 | [0.0;20.0] | 0.019 |
| IWQoL Lite – Work | 92 | 9.1 | 12.3 | <0.001 |
| IWQoL Lite - Total score | 27 | 10.5 | [4.0;23.4] | <0.001 |
| EQ-5D-3L Health Today | 92 | 6.2 | 14.7 | 0.031 |
| WEMWBS | 29 | 7.9 | 8.3 | <0.001 |
| PHQ-9 | 27 | -3.6 | 4.4 | <0.001 |
| GAD-7 | 27 | -1.0 | [-3.5;0.5] | 0.028 |
| PSQI | 31 | -2.2 | 2.3 | <0.001 |
| 6-minute walk test distance (m) | 34 | 34.5 | [16.5;50.5] | <0.001 |
| 6-minute walk test post-test heart rate (bpm) | 34 | -2.3 | 22.0 | 0.542 |
| 6-minute walk test RPE | 34 | 0.0 | [-1.5;0.0] | 0.011 |
| Sit to stand duration (s) | 34 | -0.2 | 1.9 | 0.492 |
| Right hand max (kg) | 34 | 2.18 | 4.06 | 0.004 |
| Left hand max (kg) | 34 | 1.03 | 3.89 | 0.132 |
| Dominant hand max (kg) | 34 | 2.40 | 4.16 | 0.002 |

Data presented as mean or median where appropriate with corresponding standard deviation (SD) or interquartile range (IQR). Abbreviations: BMR: basal metabolic rate, bpm: beats per minute, CoEQ: Control of Eating Questionnaire, EQ-5D-3L: EuroQol five dimensions three levels, GAD-7: Generalised Anxiety Disorder-7, ITT: intention-to-treat; IQR: interquartile range, IWQoL-Lite: Impact of Weight on Quality of Life-Lite, PHQ-9: Patient Health Questionnaire-9, PoF: Power of Food Scale, PSQI: Pittsburgh Sleep Quality Index, RPE: Rating of Perceived Exertion, SD: standard deviation, WEMWBS: Warwick Edinburgh Mental Wellbeing Scale.

### Physical activity

Changes in self-reported PA levels are reported in Supplementary Table 17 (ITT) and 18 (PP). At 6-months follow-up, there was a significant increase in vigorous PA (60 mins/week (IQR: 0.0,180.0), p = 0.018) and a significant decrease in time spent sitting (−61 mins/weekday (SD: 110), p = 0.014) from baseline. There was no significant change in total, moderate or walking PA, or total MET-minutes. Results were similar in the PP sample.

### Clinical/Patient-reported outcomes

Changes in clinical and patient-reported outcomes are reported in Table 3 (ITT) and Supplementary Table 19 (PP).

### Clinical

Percentage weight loss was significantly lower at 6-months follow-up from baseline (−3.5% (IQR: –8.6, 1.1), p < 0.001) (Table 3). Weight (−3.8 kg (IQR: –8.5,1.0), p = 0.001), BMI (−1.0 kg/m^2^ (IQR: –2.8,0.3), p = 0.001), fat mass (−2.3 kg (IQR: –5.6,0.9), p < 0.001), body fat percentage (−1.5 % (SD: 2.3), p = 0.001), visceral fat rating (−1.0 (IQR: –1.0,0.0), p = 0.001), fat-free mass (−0.8 kg (IQR: –2.6,0.2), p < 0.001), muscle mass (−0.8 kg (IQR: –2.4,0.2), p = 0.010), bone mass (0.0 kg (IQR: –0.1,0.0), p = 0.007), total body water mass (−1.0kg (IQR: – 2.1,0.0), p < 0.001), estimated basal metabolic rate (BMR) (−31kcal (IQR: –93,12), p = 0.003), systolic blood pressure (−8.1 mmHg (SD: 13.2), p = 0.001), heart rate (−5.3 beats per minute (SD: 10.1), p = 0.005), alanine transaminase (−2.0 IU/L (IQR: –4.8,0.8), p = 0.022) and c-reactive protein (−0.3 mg/l (IQR: –1.2,0.1), p = 0.017) were significantly lower at 6-months follow-up from baseline (Table 3). Total body water percentage (0.9 % (SD: 1.9), p = 0.006) and albumin (0.8 g/l (SD: 1.8), p = 0.013) were significantly higher at 6-months follow-up from baseline.

### Eating behaviour

Power of Food (−0.9 (IQR: –1.3, –0.4), p < 0.001), available (−0.8 (IQR: –1.4, –0.7), p < 0.001), present (−1.2 (SD: 1.1), p < 0.001) and tasted (−0.6 (IQR: –1.0, –0.2), p < 0.001) were significantly lower at 6-months follow-up from baseline. Control of Eating Questionnaire (CoEQ) craving control (25.0 (SD: 20.1), p < 0.001) and positive mood (8.7 (SD: 20.6), p = 0.037) were significantly higher at 6-months follow-up from baseline. CoEQ sweet craving (−12.9 (SD: 10.7), p < 0.001), savoury craving (−18.4 (SD: 17.8), p < 0.001), and craving for nominated food (−27.5 (SD: 27.7), p < 0.001) were significantly lower at 6-months follow-up from baseline (Table 3).

### Mental health and quality of life

Impact of Weight on Quality of Life-Lite (10.5 (IQR: 4.0, 23.4), p < 0.001), EuroQol five dimensions three levels health today (6.2 % (SD: 14.7), p = 0.031), Warwick Edinburgh Mental Wellbeing Scale (7.9 (SD: 8.3), p < 0.001) were significantly higher at 6-months follow-up from baseline. Patient Health Questionnaire-9 (−3.6 (SD: 4.4), p < 0.001) and Generalised Anxiety Disorder-7 (−1.0 (IQR: –3.5, 0.5), p = 0.028) were significantly lower at 6-months follow-up from baseline (Table 3).

### Sleep

Pittsburgh Sleep Quality Index (−2.2 (SD: 2.3), p < 0.001) was significantly lower at 6-months follow-up from baseline.

### Physical fitness

6-minute walk test distance walked (35 m (IQR: 17, 51), p < 0.001) and right (2.2 kg (SD: 4.1), p = 0.004) and dominant (2.4 kg (SD: 4.2), p = 0.002) maximum handgrip strength increased at 6-months follow-up from baseline. 6MWT rating of perceived exertion significantly decreased (0.0 s (IQR: –1.5, 0.0), p = 0.011). There was no significant change in sit-to-stand test duration or left maximum handgrip strength.

Changes in clinical and patient-reported outcomes were consistent in the PP sample (Supplementary Table 19 (PP)).

## Discussion

### Summary of results

This is the first study to assess the feasibility and acceptability of a BSP to reduce UPF intake in UK adults. The exploratory findings indicate high adherence, acceptability and perceived benefit to participants, with significant reductions in UPF intake. Furthermore, the intervention was associated with significant reductions in weight and improvements across several clinical and patient-reported outcomes.

### Retention/feasibility

Retention was 68%. In a systematic review of behavioural interventions targeting UPF reduction (Buck et al., retention across fifteen studies ranged from approximately 40% to 100%, with retention in most exceeding 70%, with only three studies reporting substantially lower rates^24^. Thus, the retention rate observed here sits at the lower end of most studies. However, by the time participants were offered the UPDATE BSP, they had already been in an intensive trial for 6 months. Therefore, some dropouts were likely not from the BSP itself, but from a desire to step away from the trial in general. Uptake and retention may differ when the BSP is offered as a standalone intervention.

### Intervention delivery fidelity

The intervention was feasible to deliver. Overall fidelity scores indicated proficient delivery in the introductory and first behavioural support sessions, and competent delivery in the month 2 session^35^. These suggest that core components of the BSP were delivered largely as intended, particularly in earlier stages. BCTs delivered in the initial sessions demonstrated higher delivery fidelity than BCTs introduced in later sessions. This suggests that structured and content-driven components may be easier to deliver consistently, particularly in the beginning where sessions are more standardised. As the BSP progressed, sessions became increasingly tailored to participants’ individual circumstances and goals, which may have reduced the opportunity for consistent delivery of some BCTs and contributed to greater variability in fidelity over time.

#### Acceptability

Qualitative analyses of interviews at the end of the BSP suggest that the intervention was broadly acceptable and valued by participants, though not without limitations. Acceptability appeared to be largely linked to the balance of support and flexibility provided by the one-to-one sessions, with pacing and duration deemed manageable. Perceived benefits associated with reducing UPF (e.g. improvements in wellbeing, confidence and control of overeating, and ability to sustain new habits and healthier identities) appeared to reinforce engagement with the BSP. Meanwhile, varied relevance of specific components such as the tracking booklet and group-based sessions emphasised the importance of tailoring and adaptability within multi-component interventions. Together, these findings suggest that acceptability was closely linked to the relational and reflective aspects of support, especially one-to-one sessions. This also highlighted intervention components that could be optimised to better accommodate differing preferences and support longer-term change.

These generally positive observations from interviews align with the high adherence of 88% reported in the ITT sample, and 100% adherence in nearly 60% of all starters. Acceptability of previous interventions has been mixed^24^. Most interventions have been generally acceptable to participants, with some showing strong sustained engagement^21,39^. However, some report substantial issues driven by structural barriers or dissatisfaction with the intervention itself^40,41^.

#### Changes in COM-B

Positive changes in COM-B domains were reported in exit interviews and observed from pre-post changes in the COM-B questionnaire, with significant improvements in COM-B healthy eating and PA scores, as well as specific domains linked to the BCTs used in the intervention. This included habits, action planning, action control, motivation and reinforcement. Some COM-B questionnaire domains that scored highly at baseline (>80%, e.g. knowledge, perceived behavioural control, attitude, intentions) did not significantly change by the end of the BSP, whilst some domains that scored low at baseline (<50%, e.g. action planning, action control, habit and reinforcement) did significant improve by the end of the intervention. This highlights the ceiling effect across some domains, yet capacity to address barriers. Notably, several COM-B domains with low-moderate baseline scores that did not significantly improve at the end of the intervention tended to be domains related to factors outside the control of the individual (e.g. work environment, social support, subjective norms). These provide potential domains to target with greater/wider support.

Across previous interventions, whilst behavioural improvements were commonly reported (such as reductions in UPF intake or increases in healthier food choices, typically following education, counselling, motivational strategies or skill-building activities), none explicitly assessed or reported COM-B domains, nor did they assess the underlying processes that enable or prevent behaviour change^24^. As a result, mechanisms such as planning, habit formation self-regulation, confidence, or competing external demands were not measured. In contrast, the present study explicitly measured and demonstrated improvements in several of these behavioural processes, offering insights into how and why behavioural changes occurred.

#### Change in diet

Changes in self-reported dietary intake indicated significant reductions in UPF intake at the end of the BSP, the primary target behaviour. There were concomitant reductions in total energy, free sugar, saturated fat and salt intake, and increases in MPF intake. Fibre intake decreased, but in proportion with the lower energy intake. Fruit and vegetable intake did not significantly differ. Success in reducing UPF has varied across previous studies^24^. Some interventions have resulted in notable reductions in UPF intake, including reports of almost halving energy intake from UPF or substantial decreases in daily servings^23,25^, similar to the percentage reductions reported here. Others have shown moderate improvements, such as reductions in UPF intake that did not extend to broader dietary or physiological change (e.g.^26^), and some interventions have reported limited or non-meaningful changes^40,41^. However, direct comparison across studies is challenging, with the use of different metrics and assessment methods to measure UPF intake.

#### Change in clinical / patient-reported outcomes

Compared with baseline, participants demonstrated significant body weight reductions and improvements in several other clinical and patient-reported outcomes at the end of the BSP. In previous BSPs, changes in clinical outcomes have varied considerably^24^. Some intensive, multi-component programmes (e.g.^22,23^) reported clear improvements in weight or metabolic markers, while others showed only small changes despite shifts in dietary behaviour^24^. However, whilst the findings reported here are promising, as a pre-post exploratory analysis, results must be confirmed in a future efficacy trial.

#### Implications

These results support the hypothesis that a well-designed, evidence-based behavioural intervention targeting UPF reduction in UK adults could be effective in enacting behaviour change, improving diet, PA levels, and clinical/patient-reported outcomes. An aim of the study was to further develop the UPDATE BSP based on participant feedback^29^. The preliminary results provide rationale to further test the improved BSP to assess intervention efficacy.

#### Strengths and limitations

Strengths include being the first intervention specifically designed to support UK adults to reduce their UPF intake, using evidence-based behaviour change techniques and intervention functions. Furthermore, comprehensive qualitative and quantitative data facilitated assessment of self-reported beliefs and attitudes across multiple domains of diet, exercise and lifestyle.

However, there are several limitations. First, efficacy cannot be determined in this study, as it was not statistically powered to assess changes in outcomes, and the intervention was assessed in a pre-post exploratory design with no control arm. As the intervention was offered after Stage 1, participants did not necessarily participate with the aim of participating in Stage 2, which may have influenced adherence and retention. Furthermore, users of weight loss medications were excluded (as part of Stage 1 eligibility criteria), limiting overall understanding of the intervention in the context of weight loss medications.

### Conclusion

In this first intervention designed to support UK adults to reduce their UPF intake, preliminary results indicate good feasibility and acceptability, with improvements in targeted behavioural mechanisms and health-related outcomes. These exploratory findings provide rationale to further develop and test the programme in controlled trials.

## Declarations

### Ethics approval and consent to participate

Sheffield Research Ethics Committee approved the trial (22/YH/0281).

### Consent for publication

Not applicable

### Availability of data and materials

The data used is this study is available from the corresponding author on reasonable request and following approval of data sharing agreements.

### Competing interests

SD receives royalties from Amazon for a self-published book that mentions UPF, payments from Red Pen Reviews as a contributor, consultancy work for Consensus, Mindhouse, Morgan & Morgan and Androlabs, and travel fees from a USDA National Institute of Food and Agriculture grant, (AFRI project 1033399) for a workshop on food processing classifications. RB from May 2023 is an employee and shareholder of Eli Lilly and Company. ACB declares researcher-led grants from the National Institute for Health Research, Rosetrees Trust, MRC, INNOVATE UK, British Dietetic Association, British Association of Parenteral and Enteral Nutrition, BBRSC, the Office of Health Improvement and Disparities and Novo Nordisk. ACB reports honoraria from Novo Nordisk, Lilly, Office of Health Improvement and Disparity, Johnson and Johnson and Obesity UK outside the submitted work and is on the Medical Advisory Board and shareholder of Reset Health Clinics Ltd. CAGW-K is a shareholder in Queen Square Analytics. JM reports institutional funding from Novo Nordisk, Rhythm Pharmaceuticals and Innovate UK outside the submitted work. CVT receives royalties for a book on UPF and has been paid for other broadcasting on this subject (podcast and BBC documentaries). CB, GNH, RC, FCJ, TR and AF report no conflicts of interest.

## Funding

This work was supported by National Institute for Health and Care Research Biomedical Research Centre (NIHR BRC) grant BRC530a and Rosetrees Trust grant PGL22/100041.

## Author contributions

CB: investigation, data collection, data interpretation; SD – data collection, data analysis, data interpretation; writing first manuscript draft, GNH: investigation, data collection; RC, ACB: funding; FCJ: data collection; EB: investigation; CR: data collection; TR: data collection; JM, CvT: funding; CAGW-K: funding; RB: conceptualisation, funding, supervision; AF: conceptualisation, funding, supervision, data interpretation. All authors reviewed and agreed to the final version of the manuscript.

## Acknowledgements

The authors firstly thank all participants for their time and effort as part of the study, and Dr Taylor Willmott for their support on using the COM-B questionnaire.

## Notes

### Clinical Trial

NCT05627570

### Clinical Protocols

https://bmjopen.bmj.com/content/15/10/e107435

### Author Declarations

The Yorkshire & The Humber – Sheffield Research Ethics Committee approved the trial on 22 December 2022 (22/YH/0281), and the study was prospectively registered on ClinicalTrials.gov (NCT05627570).

