## Supplementary Materials for "UPDATE trial Stage 2: a pre-post exploratory analysis of a behavioural support intervention to reduce ultra-processed food intake, increase minimally processed food intake, and increase physical activity in adults living with overweight or obesity"

**Tables**

1. **Supplementary Table 1: Behavioural support programme introductory call: TDF and intervention components**
2. **Supplementary Table 2: Behavioural support programme month 1 call: TDF and BCTs**
3. **Supplementary Table 3: Behavioural support programme month 2 call: TDF and BCTs**
4. **Supplementary Table 4: Adapted Dreyfus rating scale for the intervention BCT calls**
5. **Supplementary Table 5: Baseline characteristics of all participants commencing the behavioural support intervention**
6. **Supplementary Table 6: Baseline characteristics of participants attending the behavioural support intervention 6-month follow-up (ITT sample)**
7. **Supplementary Table 7: Baseline characteristics of participants with 100% attendance on the behavioural support intervention (PP sample)**
8. **Supplementary Table 8: Baseline characteristics of participants withdrawing or dropping out of the behavioural support intervention**
9. **Supplementary Table 9: UPDATE introductory calls BCT checklist, intended delivery technique and mean fidelity scores.**
10. **Supplementary Table 10: UPDATE month 1 calls BCT checklist, intended delivery technique and mean fidelity score.**
11. **Supplementary Table 11: UPDATE month 2 calls BCTs checklist, intended delivery technique and mean fidelity score.**
12. **Supplementary Table 12: PP: Healthy eating COM-B questionnaire scores at baseline, and changes in healthy eating COM-B questionnaire from baseline to 6-month follow-up**
13. **Supplementary Table 13: ITT: Physical activity COM-B questionnaire scores at baseline, and changes in physical activity COM-B questionnaire from baseline to 6-month follow-up**
14. **Supplementary Table 14: PP: Physical activity COM-B questionnaire scores at baseline, and changes in physical activity COM-B questionnaire from baseline to 6-month follow-up**
15. **Supplementary Table 15: ITT: Self-reported dietary intake at baseline and 6-months follow-up, and changes in self- reported dietary intake from baseline to 6-month follow-up**
16. **Supplementary Table 16: PP: Self-reported dietary intake at baseline and 6-months follow-up, and changes in self- reported dietary intake from baseline to 6-month follow-up**
17. **Supplementary Table 17: ITT: Self-reported physical activity levels at baseline and 6-months follow-up, and changes in self- reported physical activity levels from baseline to 6-month follow-up**
18. **Supplementary Table 18: PP: Self-reported physical activity levels at baseline and 6-months follow-up, and changes in self- reported physical activity levels from baseline to 6-month follow-up**
19. **Supplementary Table 19: PP: Changes in clinical and patient-reported outcomes from baseline to 6-month follow-up**

### Methods: Clinical/patient outcomes

Weight was measured using an electronic scale (Tanita DC-430MAS; Tanita), and body composition was assessed via bioelectrical impedance analysis (BIA) (Tanita) after an overnight fast. Waist circumference (WC) was measured using a tape measure at the iliac crest. BMI was derived from weight and height (in kg/m^2^), and waist-to-height ratio from WC and height. Blood pressure and heart rate were measured using an automated sphygmomanometer and pulse oximeter. Fasting glucose, HbA1c, liver function (bilirubin, alkaline phosphatase, alanine transaminase and albumin), lipids (total cholesterol, HDL-C, LDL-C, total-cholesterol-to-HDL ratio, non-HDL-C and triglycerides) and C-reactive protein were measured from venous blood samples.

Barriers and facilitators to healthy eating and PA were assessed using a 257-item questionnaire based on COM-B, adapted from Willmott et al.^1^. The questionnaire covered each COM-B domain for diet (n = 120 items) and PA (n = 137 items). Responses were averaged within each domain and combined into a summary score for diet and PA for each participant. The severity and type of food cravings was assessed using the Control of Eating Questionnaire (CoEQ)^2^. The psychological impact of living in food-abundant environments was assessed using the Power of Food (PoF) scale^3^. Mental wellbeing was assessed using the Warwick-Edinburgh mental well-being scale (WEMWBS)^4^, severity of depression symptoms was assessed using the 9-item Patient Health Questionnaire (PHQ-9)^5^, severity of anxiety symptoms was assessed using the 7-item Generalised Anxiety Disorder assessment (GAD-7)^6^. Quality of life was assessed using EuroQol 5-Dimensions 3-Levels (EQ-5D-3L)^7^. Weight-related quality of life was assessed using the Impact of Weight on Quality of Life-Lite (IWQOL-Lite)^8^. Sleep was assessed using the Pittsburgh Sleep Quality Index (PSQI)^9^, where a higher score indicates more disturbed sleep.

Physical function was assessed using the six-minute walk test (6MWT)^10^, sit-to-stand test^11^, and handgrip strength test (using a handgrip dynamometer (Jamar Hydraulic Hand Dynamometer, Patterson Medical)).

**Behavioural support programme: session-specific behaviour change technique (BCT) checklists**

**Supplementary Table 1: Behavioural support programme introductory call: TDF and intervention components**

| **Introductory call** | **Intervention Component** | **Behaviour Change Techniques**  **BCTTv1** | **No.** | **TDF** |
| --- | --- | --- | --- | --- |
| **Expertise and support** | Introductions: introduce self as behavioural scientist working with the trial team at UCL | Credible source | 9.1 | SOCIAL INFLUENCES |
|  | Ask why they originally agreed to take part in UPDATE | Review behaviour goal | 1.5 | GOALS |
|  | Tell them we will send notes after the call because some people find it helpful to have records | Prompts/cues | 7.1 | SOCIAL INFLUENCES |
| **Exploring eating behaviour before trial started** | Review current eating behaviour (habits/preferences) and identify drivers for unhealthy/high UPF diet  Identify discrepancies between current behaviour and recommended diet  Problem-solve factors influencing eating behaviour | Self-monitoring of behaviour  Problem solving  Discrepancy between current behaviour and goal | 2.3  1.2  1.6 | BEHAVIOURAL REGULATION |
| **Awareness of UPF** | Explaining that one diet was high in UPF | Information about health consequences | 5.1 | KNOWLEDGE |
| **Self-monitoring** | Ask participant to record food/beverage intake over the next week | Self-monitoring of behaviour | 2.3 | BEHAVIOURAL REGULATION |
| **Support** | Offer group sessions focused on sharing experiences, encouragement and emotional support with peers  Offer group sessions for exchanging practical tips and resources with other participants | Social support (emotional) | 3.3 | SOCIAL INFLUENCES |
|  |  | Social support (practical) | 3.2 |  |

*Abbreviations: TDF: Theoretical Domains Framework; UPF: ultra-processed food.*

**Supplementary Table 2: Behavioural support programme month 1 call: TDF and BCTs**

| **Month 1 call** | **Intervention Component** | **Behaviour Change Techniques**  **BCTTv1** | **No.** | **TDF** |
| --- | --- | --- | --- | --- |
| **Expertise and support** | Introductions: introduce self as behavioural scientist working with the trial team at UCL | Credible source | 9.1 | SOCIAL INFLUENCES |
|  | Tell them we will send notes after the call because some people find it helpful to have records | Prompts/cues | 7.1 | SOCIAL INFLUENCES |
| **Booklet/website** | Introduce the booklet/website | Adding objects to the environment | 12.5 | ENVIRONMENTAL CONTEXT |
| **Awareness of UPF prior to behavioural support/education** | Problem solve barriers to recognising UPF | Problem solving | 1.2 | BELIEFS ABOUT CAPABILITIES |
| **UPF scientific evidence** | Introduce the science about UPF (Kevin Hall + other evidence) | Information about health consequences  Credible source | 5.1  9.1 | KNOWLEDGE  SOCIAL INFLUENCES  BELIEFS ABOUT CONSEQUENCES |
| **Nova classification** | Introducing Nova; endorsed by World Cancer Research Fund | Credible source | 9.1 | SOCIAL INFLUENCES |
|  | What are UPFs | Information about health consequences | 5.1 | KNOWLEDGE  BELIEF ABOUT CONSEQUENCES |
|  | How to recognise UPF | Instruction on how to perform the behaviour | 4.1 | SKILLS |
|  | Information about links between UPF and health | Information about health consequences | 5.1 | KNOWLEDGE  BELIEF ABOUT CONSEQUENCES |
| **Checking to see if understanding and confidence has grown after education components** | Assess confidence in recognising UPF and identify barriers  Problem-solve ways to increase confidence  Discuss what they have learned about UPF and it’s impact | Problem solving  Information about health consequences | 1.2  5.1 | BELIEFS ABOUT CAPABILITIES  KNOWLEDGE |
| **Feedback on diet prior to UPDATE participation** | Tell participant how much of their diet is made up of UPF  Feedback on last seven days food/beverage diary  Ask participant where they think the UPF comes from in their current diet  Identify their current dietary approach /pattern, how they shop, cook etc | Feedback on behaviour  Discrepancy between current behaviour and goal  Information about health consequences | 2.2  1.6  5.1 | BEHAVIOURAL REGULATION  BELIEFS ABOUT CONSEQUENCES  SKILLS & SOCIAL INFLUENCES |
| **Notes and Reflections** | Review recent eating behaviour and identify drivers for high UPF consumption  Provide feedback on their behaviour | Self-monitoring of behaviour  Feedback on behaviour | 2.3  2.2 | BEHAVIOURAL REGULATION |
| **Instruction on how to reduce UPF** | Discuss benefits of reducing the amount of UPF in their diet and increasing MPF /unprocessed food | Information about health consequences | 5.1 | BELIEFS ABOUT CONSEQUENCES |
|  | Introduce how to avoid UPF | Problem solving | 1.2 | BELIEFS ABOUT CAPABILITIES |
|  | Discuss motivation to reduce UPF | Pros & cons | 9.2 | MOTIVATIONS & GOALS |
|  | Discuss barriers to reducing UPF and brainstorm solutions | Problem Solving | 1.2 | ENVIRONMENTAL CONTEXT |
| **Goal setting/action planning (behaviour)** | Discuss goal(s) defined in terms of the behaviour to be achieved | Goal setting (behaviour) | 1.2 | BEHAVIOURAL REGULATION |
|  | Promote goal setting resource in booklet/website | Adding objects to the environment | 12.5 | ENVIRONMENTAL CONTEXT |
|  | Discuss specific plans and introduce planning/ rate confidence form to help ensure goals are achieved | Action planning  Adding objects to the environment | 1.4  12.5 | GOALS  ENVIRONMENTAL CONTEXT |
| **Booklet/website resources: Recipes, snack swaps, mapping of local area for food outlets** | Go through resources such as recommendations for recipes, cooking from scratch, local food environment availability etc | Problem solving  Instructions on how to perform the behaviour | 1.2  4.1 | ENVIRONMENTAL CONTEXT  KNOWLEDGE |
| **Habits** | Describe habit formation through repetition of a behaviour e.g. looking regularly at goal/action planner | Behavioural practice/rehearsal  Habit formation | 8.1  8.3 | SKILLS |
| **Self-monitoring** | Explain purpose of self-monitoring and introduce tracker | Self-monitoring of behaviour | 2.3 | BEHAVIOURAL REGULATION |
| **Goal setting (outcome** | Discuss key goal in terms of a positive outcome of wanted behaviour | Goal setting (outcome) | 1.3 | GOALS |
| **Support** | Promote asking for emotional encouragement from friend/colleague/family member | Social support (emotional) | 3.3  3.2 | SOCIAL INFLUENCES |
|  | Promote asking for practical help from friends/colleague/family member | Social support (practical) |  |  |

*Abbreviations: BCT: Behaviour change technique; TDF: Theoretical Domains Framework; UPF: ultra-processed food.*

**Supplementary Table 3: Behavioural support programme month 2 call: TDF and BCTs**

| **Month 2 call** | **Intervention Component** | **Behaviour Change Techniques**  **BCTTv1** | **No.** | **TDF** |
| --- | --- | --- | --- | --- |
| **Checking understanding and knowledge** | Identify issues recognising UPF and find ways to improve awareness  Practice identifying UPF | Problem solving  Behavioural practice/rehearsal | 1.2  8.1 | BELIEFS ABOUT CAPABILITIES  SKILLS |
| **Goal map** | Have they designed a goal map | Review outcome goal | 1.7 | GOALS |
| **Provide feedback** | Provide feedback on goal map | Feedback on behaviour | 2.2 | BEHAVIOURAL REGULATION |
| **Assess success this month** | Have you reduced the amount of UPF in your diet | Review behaviour goal | 1.5 | GOALS |
| **Goal setting/Action planning** | Remind them of their goals and plans from last month  Thoughts and feelings about goal set and action plan set last month  Ask how they are getting on with their goals and plans | Review behaviour goals  Discrepancy between current behaviour and goal | 1.5  1.6 | GOALS |
| **Self-monitoring** | Ask them if they are self-monitoring/tracking activity | Self-monitoring of behaviour | 2.3 | BEHAVIOURAL REGULATION |
| **Habit** | Ask them if they are creating habits by looking regularly at goal/action plan and tracker | Behavioural practice/rehearsal  Habit formation | 8.1  8.3 | KNOWLEDGE |
| **Provide feedback** | Feedback on goal setting/action planning and activity so far; provide encouragement and enthusiasm for continued adherence to programme; monitor and provide informative or evaluative feedback on performance of the behaviour e.g. reduced number of UPF convenience foods, cooked more from scratch | Feedback on behaviour | 2.2 | BEHAVIOURAL REGULATION |
| **Set new goal/action plan for next month** | Ask if they want to change their goals and plans for next month  Analyse or prompt the participant to analyse factors influencing the behaviours and generate strategies that include overcoming the barriers and/or increasing facilitators | Goal setting (behaviour)  Action planning  Problem solving | 1.1  1.4  1.2 | GOALS  BEHAVIOURAL REGULATION |
| **Repeat any of the points from Month 1 as appropriate** |  |  |  |  |

*Abbreviations: BCT: Behaviour change technique; TDF: Theoretical Domains Framework; UPF: ultra-processed food.*

| **Scoring** | **Examples** | **Delivery Fidelity Category** |
| --- | --- | --- |
| **0** | **Absence of feature and/ or highly inappropriate performance** | **Low fidelity** |
| **1** | **Minimal use of feature and/ or inappropriate performance** | **Low fidelity** |
| **2** | **Scope for improvement, alongside numerous minor and some major inconsistencies** | **Scope for improvement** |
| **3** | **Competent, good features but some minor inconsistencies or problems** | **Competent** |
| **4** | **Very good features, few inconsistencies or problems** | **Proficient** |
| **5** | **Excellent features, no problems or inconsistencies** | **Expert** |

**Supplementary Table 4: Adapted Dreyfus rating scale for the intervention BCT calls (Cross et al., 2022)**

*From: Cross R, Greaves CJ, Withall J, Rejeski WJ, Stathi A. Delivery fidelity of the REACT (REtirement in ACTion) physical activity and behaviour maintenance intervention for community dwelling older people with mobility limitations. BMC Public Health 2022; 22: 1112. Abbreviations: BCT: Behaviour change technique.*

**Supplementary Table 5: Baseline characteristics of all participants commencing the behavioural support intervention**

| **BSP Starters, N = 41** | **Overall** |
| --- | --- |
| Age at screening (years) | 43.2 (11.4) |
| Sex (Female) | 38 (92.7%) |
| Ethnicity (White) | 30 (73.2%) |
| Occupation |  |
| Doctor | 1 (2.4%) |
| Nurse | 17 (41.5%) |
| AHP | 8 (19.5%) |
| Management | 3 (7.3%) |
| Administrative | 7 (17.1%) |
| Other | 5 (12.2%) |
| Night shift worker | 5 (12.2%) |
| Education level |  |
| None | 0 (0.0%) |
| GCSE/O-level equivalent | 3 (7.3%) |
| A level or equivalent | 3 (7.3%) |
| Degree | 17 (41.5%) |
| Postgraduate | 18 (43.9%) |
| Other | 0 (0.0%) |
| Marital status |  |
| Single | 16 (39.0%) |
| Married | 12 (29.3%) |
| Living together | 3 (7.3%) |
| Separated | 2 (4.9%) |
| Divorced | 4 (9.8%) |
| Widowed | 4 (9.8%) |
| Civil partnership | 0 (0.0%) |
| Family history of obesity | 24 (58.5%) |
| Family history of diabetes | 16 (39.0%) |
| Family history of cardiovascular disease | 16 (39.0%) |
| Smoking status |  |
| Yes, current | 4 (9.8%) |
| Yes past | 13 (31.7%) |
| No, never | 24 (58.5%) |
| AUDIT-C | 2.7 (1.5) |
| Weekly units of alcohol | 2.0 [1.0;4.0] |
| Weight (kg) | 90.36 (12.81) |
| Height (m) | 1.66 (0.07) |
| Body mass index (kg/m^2^) | 32.6 (4.0) |
| 25-29.9 kg/m^2^ | 14 (34.15%) |
| 30-34.9 kg/m^2^ | 16 (39.02%) |
| 35-39.9 kg/m^2^ | 11 (26.83%) |
| Waist circumference (cm) | 97.18 (10.36) |
| Waist-to-height ratio | 0.58 (0.06) |
| Fat mass (kg) | 34.50 (8.26) |
| Fat-free mass (kg) | 55.86 (6.48) |
| Systolic blood pressure (mmHg) | 129.56 (14.43) |
| Diastolic blood pressure (mmHg) | 73.60 (8.44) |
| Resting heart rate (bpm) | 74.39 (9.90) |
| HbA1c (%) | 5.51 (0.42) |
| Fasted glucose (mmol/L) | 4.85 [4.60;5.20] |
| Total cholesterol (mmol/L) | 5.22 (0.87) |

*Data presented as mean, count or median where appropriate with corresponding standard deviation, percentage, or interquartile range, respectively, in brackets. Abbreviations: AHP, allied health professional; AUDIT-C, alcohol use disorders identification test–consumption.*

**Supplementary Table 6: Baseline characteristics of participants attending the behavioural support intervention 6-month follow-up (ITT sample)**

| **ITT, N = 34** | **Overall** |
| --- | --- |
| BSP 1-1 session attendance | 88.6% (20.2) |
| Age at screening (years) | 44.7 (11.3) |
| Sex (Female) | 32 (94.1%) |
| Ethnicity (White) | 25 (73.5%) |
| Occupation |  |
| Doctor | 0 (0.0%) |
| Nurse | 13 (38.2%) |
| AHP | 7 (20.6%) |
| Management | 3 (8.8%) |
| Administrative | 7 (20.6%) |
| Other | 4 (11.8%) |
| Night shift worker | 4 (11.8%) |
| Education level |  |
| None | 0 (0.0%) |
| GCSE/O-level equivalent | 3 (8.8%) |
| A level or equivalent | 3 (8.8%) |
| Degree | 14 (41.2%) |
| Postgraduate | 14 (41.2%) |
| Other | 0 (0.0%) |
| Marital status |  |
| Single | 13 (38.2%) |
| Married | 9 (26.5%) |
| Living together | 3 (8.8%) |
| Separated | 2 (5.9%) |
| Divorced | 4 (11.8%) |
| Widowed | 3 (8.8%) |
| Civil partnership | 0 (0.0%) |
| Family history of obesity | 19 (55.9%) |
| Family history of diabetes | 13 (38.2%) |
| Family history of cardiovascular disease | 14 (41.2%) |
| Smoking status |  |
| Yes, current | 4 (11.8%) |
| Yes past | 10 (29.4%) |
| No, never | 20 (58.8%) |
| AUDIT-C | 2.7 (1.5) |
| Weekly units of alcohol | 2.0 [1.0;5.5] |
| Weight (kg) | 89.36 (11.60) |
| Height (m) | 1.66 (0.06) |
| Body mass index (kg/m^2^) | 32.5 (3.7) |
| 25-29.9 kg/m^2^ | 11 (32.35%) |
| 30-34.9 kg/m^2^ | 15 (44.12%) |
| 35-39.9 kg/m^2^ | 8 (23.53%) |
| Waist circumference (cm) | 97.06 (10.64) |
| Waist-to-height ratio | 0.59 (0.07) |
| Fat mass (kg) | 33.96 (7.01) |
| Fat-free mass (kg) | 55.40 (6.04) |
| Systolic blood pressure (mmHg) | 130.07 (15.43) |
| Diastolic blood pressure (mmHg) | 73.35 (8.82) |
| Resting heart rate (bpm) | 74.38 (9.99) |
| HbA1c (%) | 5.50 (0.44) |
| Fasted glucose (mmol/L) | 4.80 [4.60;5.20] |
| Total cholesterol (mmol/L) | 5.30 (0.88) |

*Data presented as mean, count or median where appropriate with corresponding standard deviation, percentage, or interquartile range, respectively, in brackets. Abbreviations: AHP, allied health professional; AUDIT-C, alcohol use disorders identification test–consumption; ITT: intention-to-treat.*

**Supplementary Table 7: Baseline characteristics of participants with 100% attendance on the behavioural support intervention (PP sample)**

| **PP, N = 23** | **Overall** |
| --- | --- |
| BSP 1-1 session attendance | 100% |
| Age at screening (years) | 43.9 (11.1) |
| Sex (Female) | 22 (95.7%) |
| Ethnicity (White) | 17 (73.9%) |
| Occupation |  |
| Doctor | 0 (0.0%) |
| Nurse | 9 (39.1%) |
| AHP | 6 (26.1%) |
| Management | 2 (8.7%) |
| Administrative | 3 (13.0%) |
| Other | 3 (13.0%) |
| Night shift worker | 1 (4.3%) |
| Education level |  |
| None | 0 (0.0%) |
| GCSE/O-level equivalent | 1 (4.3%) |
| A level or equivalent | 2 (8.7%) |
| Degree | 8 (34.8%) |
| Postgraduate | 12 (52.2%) |
| Other | 0 (0.0%) |
| Marital status |  |
| Single | 8 (34.8%) |
| Married | 7 (30.4%) |
| Living together | 1 (4.3%) |
| Separated | 2 (8.7%) |
| Divorced | 2 (8.7%) |
| Widowed | 3 (13.0%) |
| Civil partnership | 0 (0.0%) |
| Family history of obesity | 13 (56.5%) |
| Family history of diabetes | 9 (39.1%) |
| Family history of cardiovascular disease | 9 (39.1%) |
| Smoking status |  |
| Yes, current | 1 (4.3%) |
| Yes past | 6 (26.1%) |
| No, never | 16 (69.6%) |
| AUDIT-C | 2.0 [1.0;3.0] |
| Weekly units of alcohol | 2.0 [0.5;5.0] |
| Weight (kg) | 89.67 (12.72) |
| Height (m) | 1.66 (0.06) |
| Body mass index (kg/m^2^) | 32.5 (3.6) |
| 25-29.9 kg/m^2^ | 8 (34.78%) |
| 30-34.9 kg/m^2^ | 9 (39.13%) |
| 35-39.9 kg/m^2^ | 6 (26.09%) |
| Waist circumference (cm) | 96.00 (11.56) |
| Waist-to-height ratio | 0.58 (0.07) |
| Fat mass (kg) | 34.26 (7.74) |
| Fat-free mass (kg) | 55.41 (5.74) |
| Systolic blood pressure (mmHg) | 129.07 (16.04) |
| Diastolic blood pressure (mmHg) | 71.78 (9.07) |
| Resting heart rate (bpm) | 74.39 (11.03) |
| HbA1c (%) | 5.44 (0.40) |
| Fasted glucose (mmol/L) | 4.80 [4.60;5.07] |
| Total cholesterol (mmol/L) | 5.33 (0.95) |

*Data presented as mean, count or median where appropriate with corresponding standard deviation, percentage, or interquartile range, respectively, in brackets. Abbreviations: AHP, allied health professional; AUDIT-C, alcohol use disorders identification test–consumption; PP: per protocol.*

**Supplementary Table 8: Baseline characteristics of participants withdrawing or dropping out of the behavioural support intervention**

| **Dropouts and withdrawals, N = 13** | **Dropouts and Withdrawals (N = 13)** | **Dropouts only (N = 7)** |
| --- | --- | --- |
| Age at screening (years) | 41.8 (12.5) | 36.1 (10.0) |
| Sex (Female) | 12 (92.3%) | 6 (85.7%) |
| Ethnicity (White) | 10 (76.9%) | 5 (71.4%) |
| Occupation |  |  |
| Doctor | 1 (7.7%) | 1 (14.3%) |
| Nurse | 7 (53.8%) | 4 (57.1%) |
| AHP | 1 (7.7%) | 1 (14.3%) |
| Management | 1 (7.7%) | 0 (0%) |
| Administrative | 2 (15.4%) | 0 (0%) |
| Other | 1 (7.7%) | 1 (14.3%) |
| Night shift worker | 3 (23.1%) | 1 (14.3%) |
| Education level |  |  |
| None | 0 (0%) | 0 (0%) |
| GCSE/O-level equivalent | 2 (15.4%) | 0 (0%) |
| A level or equivalent | 1 (7.7%) | 0 (0%) |
| Degree | 5 (38.5%) | 3 (42.9%) |
| Postgraduate | 5 (38.5%) | 4 (57.1%) |
| Other | 0 (0.0%) | 0 (0.0%) |
| Weight (kg) | 89.9 (15.4) | 95.2 (17.9) |
| Body mass index (kg/m^2^) | 32.4 (5.1) | 33.2 (5.5) |
| 25-29.9 kg/m^2^ | 6 (46.15%) | 3 (42.86%) |
| 30-34.9 kg/m^2^ | 3 (23.08%) | 1 (14.29%) |
| 35-39.9 kg/m^2^ | 4 (30.77%) | 3 (42.86%) |

*Data presented as mean, count or median where appropriate with corresponding standard deviation, percentage, or interquartile range, respectively, in brackets. Abbreviations: AHP, allied health professional; AUDIT-C, alcohol use disorders identification test–consumption.*

**Supplementary Table 9: UPDATE introductory calls BCT checklist, intended delivery technique and mean fidelity scores. N = 7.**

| **Intervention Behaviour Change Technique** | **Intended delivery of techniques** | **Mean Delivery Fidelity Score (SD)** |
| --- | --- | --- |
| Credible Source | Introductions: introduce self as behavioural scientist working with the trial team at UCL | 5.00 |
| Review behaviour goal | Ask why they originally agreed to take part in UPDATE | 5.00 |
| Self-monitoring of behaviour | Ask participant to record food/beverage intake over the next week | 5.00 |
| Self-monitoring of behaviour | Review eating habits/preferences and drivers of UPF intake | 4.57 |
| Discrepancy between current behaviour and goal | Identify discrepancies between current eating behaviour and recommended diet | 4.57 |
| Information about health consequences | Explaining that one diet was high in UPF | 4.86 |
| Social support (emotional) | Offer group sessions focused on sharing experiences, encouragement and emotional support with peers | 5.00 |
| Social support (practical) | Offer group sessions for exchanging practical tips and resources with other participants | 5.00 |

*Abbreviations: BCT: Behaviour change technique; UPF: ultra-processed food.*

NB Not all intended BCTs were assessed for fidelity and therefore are not included in this table.

**Supplementary Table 10: UPDATE month 1 calls BCT checklist, intended delivery technique and mean fidelity score. N = 7.**

| **Intervention Behaviour Change Techniques** | **Intended delivery of techniques** | **Mean Delivery Fidelity Score (SD)** |
| --- | --- | --- |
| Credible source  Information about health consequences | Introductions: introduce self as behavioural scientist working with the trial team at UCL  Introduce: The science of UPF (Kevin Hall’s UPF trial and other evidence)  Introducing Nova: endorsed by World Cancer Research Fund | 4.29 |
| Adding objects to the environment | Introduce the booklet/website  Promote goal setting resource in booklet/website | 5.00 |
| Problem-solving | Assess confidence in recognising UPF and identify barriers  Problem-solve ways to increase confidence  Discuss barriers to reducing UPF and brainstorm solutions | 4.43 |
| Information about health consequences | Introduce: The science of UPF (Kevin Hall + other evidence)  What UPFs are  Information about links between UPF and health  Discuss benefits of reducing the amount of UPF in their diet and increasing MPF /unprocessed food | 4.57 |
| Instruction on how to perform a behaviour | How to recognise UPF  Go through resources such as recommendations for recipes, cooking from scratch, local food environment availability etc | 4.86 |
| Information about health consequences | What have they learned so far about UPF and their impact | 4.29 |
| Feedback on behaviour | Tell participant how much of their diet is made up of UPF  Feedback on last seven days food/beverage diary | 4.71 |
| Discrepancy between current behaviour and goal | Ask participant where they think the UPF comes from in their current diet  Identify their current dietary approach /pattern, how they shop, cook, plan | 3.86 |
| Self-monitoring of behaviour | What are the drivers/motivations for eating a diet high in UPF  Explain purpose of self-monitoring and introduce tracker | 4.86 |
| Pros and cons | Discuss motivation to reduce UPF | 4.71 |
| Goal setting (behaviour) | Discuss goal(s) defined in terms of the behaviour to be achieved | 4.86 |
| Action planning | Discuss specific plans and introduce planning/ rate confidence form to help ensure goals are achieved | 4.30 |
| Behavioural practice/rehearsal | Describe habit formation through repetition of a behaviour e.g. looking regularly at goal/action planner | 4.30 |
| Habit formation | Describe habit formation through repetition of a behaviour e.g. looking regularly at goal/action planner | 4.30 |
| Goal setting (outcome) | Discuss key goal in terms of a positive outcome of wanted behaviour | 2.86 |
| Social support (emotional) | Promote asking for emotional support (e.g., encouragement, reassurance etc.) from friend/colleague/family member | 4.00 |
| Social support (practical) | Promote asking for practical support (e.g., help with tasks, problem-solving, reminders etc.) from friend/colleague/family member | 4.00 |

*Abbreviations: BCT: Behaviour change technique; UPF: ultra-processed food.*

**Supplementary Table 11: UPDATE month 2 calls BCTs checklist, intended delivery technique and mean fidelity score. N=6 calls.**

| **Intervention Behaviour Change Techniques** | **Intended delivery of techniques** | **Mean Delivery Fidelity Score (SD)** |
| --- | --- | --- |
| Problem solving | What are the benefits of reducing UPF from your diet?  Check confidence at ability to recognise UPF  Ask if they want to change their goals and plans for next month  Analyse or prompt the participant to analyse factors influencing the behaviours and generate strategies that include overcoming the barriers and/or increasing facilitators | 3.83 |
| Behavioural practice/rehearsal | What are the benefits of reducing UPF from your diet  Check confidence at ability to recognise UPF | 2.83 |
| Review outcome goal | Have they designed a goal map | 4.16 |
| Feedback on behaviour | Provide feedback on goal map | 1.67 |
| Review behaviour goal | Have they reduced the amount of UPF in their diet?  Remind them of their goals and plans from last month  Ask if they want to change their goals and plans for next month | 4.50 |
| Discrepancy between current behaviour and goal | Thoughts and feelings about goal set and action plan set last month  Ask how they are getting on with their goals and plans | 3.50 |
| Self-monitoring of behaviour | Ask them if they are self-monitoring/tracking activity | 4.60 |
| Habit formation | Ask them if they are creating habits by looking regularly at goal/action plan and tracker | 1.33 |
| Feedback on behaviour | Feedback on goal setting/action planning and activity so far; provide encouragement and enthusiasm for continued adherence to programme; monitor and provide informative or evaluative feedback on performance of the behaviour eg. reduced number of UPF convenience foods, cooked more from scratch | 3.83 |

*Abbreviations: BCT: Behaviour change technique; UPF: ultra-processed food.*

**Supplementary Table 12: PP: Healthy eating COM-B questionnaire scores at baseline, and changes in healthy eating COM-B questionnaire from baseline to 6-month follow-up**

| **Healthy eating (%), PP, N = 23** | **Baseline (N = 23)** | **Change from baseline** | | | |
| --- | --- | --- | --- | --- | --- |
|  | **Mean (SD) / Median (IQR)** | **N** | **Mean / Median** | **SD / IQR** | **P-value** |
| Total score | 61.7 (8.2) | 16 | 8.6 | 9.4 | 0.002 |
| Knowledge | 80.0 [72.3;83.8] | 17 | 2.9 | 6.4 | 0.081 |
| Decision making | 71.0 (10.5) | 17 | 1.6 | 10.2 | 0.534 |
| Decision making (pros) | 86.1 (9.4) | 17 | -3.9 | 12.4 | 0.212 |
| Decision making (cons) | 55.9 (20.0) | 17 | 7.1 | 15.3 | 0.075 |
| Action control | 41.9 (24.4) | 17 | 26.0 | 23.5 | <0.001 |
| Action planning | 33.5 (23.2) | 17 | 29.9 | 32.3 | 0.002 |
| Habit | 36.7 (19.5) | 17 | 21.4 | 22.9 | <0.001 |
| Skills | 83.3 [56.0;96.4] | 17 | 4.3 | 10.1 | 0.096 |
| Environment | 56.3 (15.6) | 17 | 11.3 | 12.2 | 0.002 |
| Environment (work) | 39.4 (25.8) | 17 | 6.2 | 20.5 | 0.230 |
| Environment (shop) | 72.2 [55.6;80.6] | 17 | 9.5 | 25.5 | 0.145 |
| Environment (home) | 62.0 (21.9) | 17 | 18.1 | 27.5 | 0.015 |
| Subjective norms | 60.4 (14.3) | 17 | -2.9 | 14.9 | 0.428 |
| Subjective norms (injunctive) | 67.4 (21.5) | 17 | -4.2 | 22.8 | 0.453 |
| Subjective norms (descriptive) | 50.0 [44.4;66.7] | 17 | -1.6 | 18.9 | 0.726 |
| Social support | 50.0 [45.0;57.5] | 17 | 0.3 | 15.5 | 0.939 |
| Social support (positive) | 37.4 (30.4) | 17 | 5.3 | 22.7 | 0.350 |
| Social support (negative) | 68.0 (21.2) | 17 | -4.7 | 25.3 | 0.454 |
| Self-efficacy | 73.2 (18.2) | 17 | 3.3 | 17.7 | 0.457 |
| Perceived behavioural control | 88.9 [66.7;100.0] | 17 | -0.3 | 15.9 | 0.933 |
| Attitude | 82.0 (13.8) | 16 | 2.6 | 8.0 | 0.213 |
| Attitude (instrumental) | 100.0 [97.2;100.0] | 16 | -2.4 | 9.1 | 0.343 |
| Attitude (affective) | 67.9 (23.6) | 16 | 7.6 | 18.0 | 0.111 |
| Goals | 59.5 (17.3) | 17 | 6.5 | 23.0 | 0.263 |
| Goals (eating) | 67.8 (22.9) | 17 | 3.6 | 24.6 | 0.555 |
| Goals (drinking) | 51.3 (18.2) | 17 | 9.3 | 25.4 | 0.150 |
| Intentions | 93.3 [83.3;100.0] | 17 | -5.7 | 16.9 | 0.183 |
| Identity | 72.2 [58.3;80.6] | 17 | 9.2 | 15.2 | 0.025 |
| Affect | 63.9 (11.8) | 17 | 2.6 | 16.5 | 0.529 |
| Affect (positive) | 75.0 [50.0;75.0] | 17 | -22.1 | 28.5 | 0.006 |
| Affect (negative) | 60.9 (30.3) | 17 | 27.2 | 35.5 | 0.006 |
| Motivation | 50.0 [49.0;56.2] | 17 | 9.4 | 15.3 | 0.022 |
| Motivation (hunger) | 83.3 [75.0;83.3] | 17 | -2.0 | 21.8 | 0.715 |
| Motivation (not hunger) | 18.8 [14.6;27.1] | 17 | 14.6 | [10.4;29.2] | <0.001 |
| Reinforcement | 41.8 (24.9) | 17 | 19.1 | 22.8 | 0.003 |
| Reinforcement (control) | 25.0 [10.4;39.6] | 17 | 23.0 | 29.6 | 0.005 |
| Reinforcement (satiety) | 48.2 (28.9) | 17 | 13.2 | 29.8 | 0.085 |
| Reinforcement (mind) | 47.8 (28.3) | 17 | 20.9 | 21.7 | 0.001 |

*Data presented as mean or median where appropriate with corresponding standard deviation (SD) or interquartile range (IQR). Abbreviations: COM-B: Capability, Opportunity, and Motivation model, IQR: inter-quartile range, PP: per protocol, SD: standard deviation.*

**Supplementary Table 13: ITT: Physical activity COM-B questionnaire scores at baseline, and changes in physical activity COM-B questionnaire from baseline to 6-month follow-up**

| **Physical activity (%), ITT, N = 34** | **Baseline (N = 34)** | **Change from baseline** | | | |
| --- | --- | --- | --- | --- | --- |
|  | **Mean (SD) / Median (IQR)** | **N** | **Mean / Median** | **SD / IQR** | **P-value** |
| Total score | 54.7 (12.4) | 25 | 4.8 | 9.0 | 0.013 |
| Knowledge | 66.7 [50.0;83.3] | 26 | 0.0 | [0.0;16.7] | 0.180 |
| Decision making | 67.0 (13.1) | 26 | 1.9 | [-6.3;9.6] | 0.510 |
| Decision making (pros) | 83.3 [72.5;92.9] | 26 | -2.8 | 11.9 | 0.237 |
| Decision making (cons) | 52.9 (18.1) | 26 | 8.3 | [-2.1;18.8] | 0.130 |
| Action control | 41.5 (29.6) | 26 | 16.1 | 27.3 | 0.006 |
| Action planning | 33.3 [0;66.7] | 26 | 10.4 | 34.4 | 0.135 |
| Habit | 22.2 [8.3;58.0] | 25 | 10.4 | 20.8 | 0.019 |
| Skills | 34.2 (21.0) | 26 | 12.5 | [0.0;21.5] | 0.004 |
| Environment | 68.4 (15.1) | 26 | 5.0 | 14.1 | 0.085 |
| Resources | 71.4 [57.1;85.7] | 26 | 0.0 | [-14.0.3;0] | 0.342 |
| Subjective norms | 55.3 (18.0) | 26 | 0.3 | 20.4 | 0.949 |
| Social support | 35.3 [20.2;51.3] | 26 | 1.9 | 17.5 | 0.589 |
| Self-efficacy | 61.8 (26.1) | 26 | 0.0 | [-5.6;9.7] | 0.862 |
| Perceived behavioural control | 80.6 [61.1;94.4] | 26 | 5.6 | [0.0;15.3] | 0.167 |
| Attitude | 72.2 [58.3;88.9] | 25 | 2.8 | [-5.6;5.6] | 0.872 |
| Attitude (instrumental) | 100.0 [83.3;100.0] | 25 | 0.0 | [-11.1;0.0] | 0.220 |
| Attitude (affective) | 55.6 [29.2;83.3] | 25 | 4.4 | 24.7 | 0.377 |
| Goals | 59.8 (21.6) | 26 | -7.8 | 16.6 | 0.024 |
| Goals (social) | 42.5 (29.4) | 26 | -8.3 | 26.4 | 0.119 |
| Goals (appearance) | 77.1 [54.2;83.3] | 26 | -9.8 | 15.7 | 0.004 |
| Goals (personal) | 68.8 [54.2;83.3] | 26 | -5.3 | 19.1 | 0.170 |
| Intentions | 66.7 [47.2;83.3] | 26 | 2.1 | 24.9 | 0.665 |
| Identity | 39.8 [17.1;57.4] | 26 | 5.2 | 21.5 | 0.229 |
| Affect | 56.9 (19.0) | 26 | 5.0 | [-2.5;20.0] | 0.022 |
| Affect (positive) | 49.1 (23.2) | 26 | 9.8 | 13.9 | 0.001 |
| Affect (negative) | 67.5 [51.2;80.0] | 26 | 7.5 | [-3.8;20.0] | 0.105 |
| Reinforcement | 43.2 (22.4) | 26 | 7.2 | 16.8 | 0.039 |

*Data presented as mean or median where appropriate with corresponding standard deviation (SD) or interquartile range (IQR). Abbreviations: COM-B: Capability, Opportunity, and Motivation model, ITT: intention-to-treat, IQR: inter-quartile range, SD: standard deviation.*

**Supplementary Table 14: PP: Physical activity COM-B questionnaire scores at baseline, and changes in physical activity COM-B questionnaire from baseline to 6-month follow-up**

| **Physical activity (%), PP, N = 23** | **Baseline (N = 23)** | **Change from baseline** | | | |
| --- | --- | --- | --- | --- | --- |
|  | **Mean (SD) / Median (IQR)** | **N** | **Mean / Median** | **SD / IQR** | **P-value** |
| Total score | 55.2 (12.3) | 16 | 6.6 | 9.9 | 0.018 |
| Knowledge | 83.3 [66.7;83.3] | 17 | 7.8 | 17.8 | 0.088 |
| Decision making | 68.0 (13.0) | 17 | 0.3 | 12.1 | 0.917 |
| Decision making (pros) | 85.0 [77.5;92.5] | 17 | -5.6 | 12.5 | 0.084 |
| Decision making (cons) | 53.6 (16.6) | 17 | 6.2 | 20.5 | 0.231 |
| Action control | 43.8 (31.2) | 17 | 19.6 | 27.0 | 0.009 |
| Action planning | 37.5 [0;66.7] | 17 | 16.2 | 27.8 | 0.029 |
| Habit | 25.0 [11.1;56.2] | 16 | 12.2 | 22.6 | 0.049 |
| Skills | 31.2 (18.4) | 17 | 16.2 | 17.1 | 0.001 |
| Environment | 72.6 (13.1) | 17 | 4.3 | 11.6 | 0.146 |
| Resources | 70.8 (19.5) | 17 | 0.0 | [-14.3;0.0] | 0.440 |
| Subjective norms | 50.0 [43.3;76.7] | 17 | -1.2 | 19.3 | 0.804 |
| Social support | 33.3 [21.2;57.1] | 17 | 0.7 | 16.0 | 0.863 |
| Self-efficacy | 62.8 (27.5) | 17 | 0.0 | [-16.7;5.6] | 0.724 |
| Perceived behavioural control | 66.7 [61.1;100.0] | 17 | 5.2 | 28.7 | 0.463 |
| Attitude | 75.4 (19.1) | 16 | 2.1 | 19.3 | 0.672 |
| Attitude (instrumental) | 100.0 [83.3;100.0] | 16 | 0.0 | [-1.4;1.4] | 0.944 |
| Attitude (affective) | 61.6 (28.5) | 16 | 5.2 | 23.0 | 0.379 |
| Goals | 58.3 [47.2;68.8] | 17 | -7.3 | 18.4 | 0.123 |
| Goals (social) | 35.7 (27.6) | 17 | -6.9 | 26.4 | 0.300 |
| Goals (appearance) | 79.2 [52.1;83.3] | 17 | -8.8 | 17.0 | 0.049 |
| Goals (personal) | 70.8 [52.1;83.3] | 17 | -6.1 | 20.4 | 0.234 |
| Intentions | 65.5 (28.5) | 17 | -2.6 | 18.7 | 0.573 |
| Identity | 38.6 (27.3) | 17 | 8.1 | 21.3 | 0.138 |
| Affect | 56.0 (18.5) | 17 | 14.4 | 13.4 | <0.001 |
| Affect (positive) | 50.0 (22.1) | 17 | 13.5 | 14.4 | 0.001 |
| Affect (negative) | 62.0 (25.9) | 17 | 15.3 | 19.3 | 0.005 |
| Reinforcement | 41.0 (21.4) | 17 | 12.1 | 15.5 | 0.005 |

*Data presented as mean or median where appropriate with corresponding standard deviation (SD) or interquartile range (IQR). Abbreviations: COM-B: Capability, Opportunity, and Motivation model, IQR: inter-quartile range, PP: per protocol, SD: standard deviation.*

**Supplementary Table 15: ITT: Self-reported dietary intake at baseline and 6-months follow-up, and changes in self- reported dietary intake from baseline to 6-month follow-up**

| **ITT N = 34** | **Baseline (N = 34)** | | **6-months follow-up (N = 29)** | | **Change from baseline (N = 29)** | | | | |
| --- | --- | --- | --- | --- | --- | --- | --- | --- | --- |
|  | **Mean** | **SD** | **Mean** | **SD** | **Mean/Median** | **SD/IQR** | **Lower 95% CI** | **Upper 95% CI** | **p-value** |
| Total energy (kcal/day) | 2202.4 | 722.4 | 1401.3 | 388.6 | -822.5 | 607.8 | -1053.7 | -591.3 | <0.001 |
| MPF (kcal/day) | 488.6 | 151.6 | 646.0 | 277.9 | 148.7 | 234.0 | 59.7 | 237.7 | 0.002 |
| PCI (kcal/day) | 65.6 | 73.4 | 46.0 | 92.9 | -27.4 | 66.5 | -52.7 | -2.1 | 0.035 |
| PF (kcal/day) | 172.8 | 110.7 | 147.1 | 153.2 | -31.7 | 160.8 | -92.9 | 29.4 | 0.297 |
| UPF (kcal/day) | 1470.6 | 555.6 | 562.3 | 315.9 | -910.5 | 621.7 | -1147.0 | -674.0 | <0.001 |
| MPF (% kcal/day) | 23.4 | 5.3 | 46.3 | 15.9 | 22.6 | 16.0 | 16.5 | 28.7 | <0.001 |
| PCI (% kcal/day) | 2.9 | 2.7 | 2.8 | 4.8 | -0.4 | 3.8 | -1.8 | 1.1 | 0.625 |
| PF (% kcal/day) | 7.8 | 4.7 | 10.6 | 10.6 | 2.6 | 10.8 | -1.5 | 6.7 | 0.199 |
| UPF (% kcal/day) | 65.6 | 6.9 | 40.2 | 19.1 | -24.8 | 19.8 | -32.4 | -17.3 | <0.001 |
| Fat (% kcal/day) | 36.9 | 4.4 | 36.2 | 8.4 | -0.9 | 7.0 | -3.6 | 1.8 | 0.491 |
| Saturated Fat (% kcal/day) | 13.3 | 2.6 | 11.5 | 4.1 | -1.9 | 4.5 | -3.6 | -0.1 | 0.035 |
| Carbohydrate (% kcal/day) | 44.4 | 4.8 | 41.4 | 6.6 | -2.9 | 6.1 | -5.2 | -0.6 | 0.016 |
| Total sugar (% kcal/day) | 17.1 | 5.0 | 16.6 | 6.0 | -0.8 | 6.6 | -3.4 | 1.7 | 0.503 |
| Total free sugar (% kcal/day) | 9.6 | 4.1 | 6.1 | 3.9 | -3.6 | 4.5 | -5.3 | -1.9 | <0.001 |
| Salt (g/day) | 6.3 | 2.3 | 3.4 | 1.6 | -3.0 | 2.4 | -3.9 | -2.0 | <0.001 |
| Protein (% kcal/day) | 16.1 | 2.4 | 21.0 | 4.2 | 5.1 | 4.3 | 3.5 | 6.8 | <0.001 |
| Fibre (g/day) | 22.0 | 8.1 | 16.7 | 6.4 | -5.4 | 5.8 | -7.6 | -3.2 | <0.001 |
| Alcohol (% kcal/day) | 2.6 | 4.0 | 1.4 | 4.7 | 0.0 | [-1.7;0.0] |  |  | 0.025 |
| Red meat (g/day) | 51.3 | 35.7 | 31.7 | 38.5 | -15.8 | 45.7 | -33.2 | 1.6 | 0.073 |
| Oily fish (g/day) | 8.7 | 16.7 | 7.7 | 23.0 | 0.0 | [-7.6;0.0] |  |  | 0.224 |
| Fruit and vegetables (portions/day) | 3.8 | 1.6 | 4.3 | 2.3 | 0.3 | 2.2 | -0.5 | 1.1 | 0.444 |

*Data presented as mean or median where appropriate with corresponding standard deviation (SD) or interquartile range (IQR). Abbreviations: CI: confidence interval, ITT: intention to treat, IQR: inter-quartile range, MPF: minimally processed food, PCI: processed culinary ingredients, PF: processed food, SD: standard deviation, UPF: ultra-processed food.*

**Supplementary Table 16: PP: Self-reported dietary intake at baseline and 6-months follow-up, and changes in self- reported dietary intake from baseline to 6-month follow-up**

| **PP N = 23** | **Baseline (N = 23)** | | **6-months follow-up (N = 21)** | | **Change from baseline (N = 21)** | | | | |
| --- | --- | --- | --- | --- | --- | --- | --- | --- | --- |
|  | **Mean** | **SD** | **Mean** | **SD** | **Mean/Median** | **SD/IQR** | **Lower 95% CI** | **Upper 95% CI** | **p-value** |
| Total energy (kcal/day) | 2297.2 | 743.8 | 1365.4 | 403.3 | -913.4 | 552.7 | -1164.9 | -661.8 | <0.001 |
| MPF (kcal/day) | 512.2 | 136.8 | 687.0 | 302.9 | 180.8 | 264.4 | 60.5 | 301.1 | 0.005 |
| PCI (kcal/day) | 75.5 | 80.3 | 50.2 | 104.7 | -28.4 | 70.6 | -60.5 | 3.7 | 0.080 |
| PF (kcal/day) | 188.2 | 120.7 | 131.1 | 138.3 | -59.9 | 156.4 | -131.1 | 11.3 | 0.094 |
| UPF (kcal/day) | 1520.2 | 578.5 | 497.1 | 258.0 | -1004.7 | 593.6 | -1275.0 | -734.5 | <0.001 |
| MPF (% kcal/day) | 23.7 | 5.2 | 50.0 | 16.3 | 26.8 | [22.3;36.4] |  |  | <0.001 |
| PCI (% kcal/day) | 3.0 | 2.7 | 3.1 | 5.4 | -0.1 | 4.2 | -2.0 | 1.8 | 0.932 |
| PF (% kcal/day) | 8.1 | 4.8 | 9.6 | 8.9 | 1.3 | 10.6 | -3.5 | 6.1 | 0.590 |
| UPF (% kcal/day) | 65.1 | 7.5 | 37.3 | 19.8 | -27.6 | 21.5 | -37.3 | -17.8 | <0.001 |
| Fat (% kcal/day) | 37.3 | 4.2 | 37.2 | 8.7 | -0.3 | 7.2 | -3.5 | 3.0 | 0.873 |
| Saturated Fat (% kcal/day) | 13.3 | 2.3 | 11.7 | 4.2 | -1.5 | 4.2 | -3.4 | 0.4 | 0.121 |
| Carbohydrate (% kcal/day) | 44.0 | 5.1 | 41.4 | 7.1 | -2.7 | 6.5 | -5.7 | 0.2 | 0.067 |
| Total sugar (% kcal/day) | 16.9 | 4.4 | 15.8 | 6.1 | -0.9 | 6.7 | -4.0 | 2.1 | 0.533 |
| Total free sugar (% kcal/day) | 9.6 | 4.1 | 5.5 | 4.0 | -3.9 | 3.9 | -5.7 | -2.1 | <0.001 |
| Salt (g/day) | 6.8 | 2.4 | 3.5 | 1.8 | -3.3 | 2.5 | -4.5 | -2.2 | <0.001 |
| Protein (% kcal/day) | 16.3 | 2.5 | 20.8 | 3.6 | 4.8 | 4.0 | 3.0 | 6.6 | <0.001 |
| Fibre (g/day) | 22.6 | 9.0 | 17.5 | 7.1 | -5.7 | 6.1 | -8.5 | -2.9 | <0.001 |
| Alcohol (% kcal/day) | 2.4 | 3.4 | 0.6 | 2.0 | -0.1 | [-2.1;0.0] |  |  | 0.004 |
| Red meat (g/day) | 53.7 | 39.2 | 38.1 | 42.0 | -11.1 | 52.2 | -34.8 | 12.7 | 0.342 |
| Oily fish (g/day) | 6.2 | 12.9 | 10.2 | 26.7 | 0.0 | [-3.1;0.0] |  |  | 0.726 |
| Fruit and vegetables (portions/day) | 4.0 | 1.6 | 4.4 | 2.5 | 0.2 | 2.4 | -0.8 | 1.3 | 0.643 |

*Data presented as mean or median where appropriate with corresponding standard deviation (SD) or interquartile range (IQR). Abbreviations: CI: confidence interval, IQR: inter-quartile range, MPF: minimally processed food, PCI: processed culinary ingredients, PF: processed food, PP: per protocol, SD: standard deviation, UPF: ultra-processed food.*

**Supplementary Table 17: ITT: Self-reported physical activity levels at baseline and 6-months follow-up, and changes in self- reported physical activity levels from baseline to 6-month follow-up**

| **ITT N = 34** | **Baseline** | | | **6-months follow-up** | | | **Change from baseline** | | | |
| --- | --- | --- | --- | --- | --- | --- | --- | --- | --- | --- |
|  | **N** | **Mean / Median** | **SD / IQR** | **N** | **Mean / Median** | **SD / IQR** | **N** | **Mean / Median** | **SD / IQR** | **p-value** |
| Total physical activity (mins/week) | 29 | 420.0 | [360.0;870.0] | 24 | 600.0 | [360.0;1125.0] | 22 | 46.6 | 657.8 | 0.743 |
| Vigorous physical activity (mins/week) | 32 | 0.0 | [0.0;60.0] | 29 | 120.0 | [0.0;300.0] | 27 | 60.0 | [0.0;180.0] | 0.018 |
| Moderate physical activity (mins/week) | 33 | 0.0 | [0.0;80.0] | 28 | 35.0 | [0;195.0] | 27 | 0.0 | [0.0;95.0] | 0.381 |
| Walking physical activity (mins/week) | 30 | 420.0 | [266.8;630.0] | 24 | 420.0 | [180.0;682.5] | 23 | -41.7 | 545.3 | 0.587 |
| Sitting (mins/weekday) | 28 | 480.0 | [300.0;555.0] | 25 | 360.0 | [300.0;450.0] | 23 | -61.1 | 110.2 | 0.014 |
| Total MET-minutes | 29 | 1386.0 | [1188.0;2892.0] | 24 | 2106.0 | [1272.0;4032.8] | 22 | 225.8 | 2305.0 | 0.651 |

*Data presented as mean or median where appropriate with corresponding standard deviation (SD) or interquartile range (IQR). Abbreviations: ITT: intention to treat, IQR: inter-quartile range, SD: standard deviation.*

**Supplementary Table 18: PP: Self-reported physical activity levels at baseline and 6-months follow-up, and changes in self- reported physical activity levels from baseline to 6-month follow-up**

| **PP N = 23** | **Baseline** | | | **6-months follow-up** | | | **Change from baseline** | | | |
| --- | --- | --- | --- | --- | --- | --- | --- | --- | --- | --- |
|  | **N** | **Mean / Median** | **SD / IQR** | **N** | **Mean / Median** | **SD / IQR** | **N** | **Mean / Median** | **SD / IQR** | **p-value** |
| Total physical activity (mins/week) | 19 | 420.0 | [318.5;1125.0] | 16 | 690.0 | [322.5;1552.5] | 14 | 104.6 | 740.7 | 0.606 |
| Vigorous physical activity (mins/week) | 22 | 0.0 | [0.0;82.5] | 19 | 135.0 | [0.0;382.5] | 18 | 65.0 | [0.0;326.2] | 0.039 |
| Moderate physical activity (mins/week) | 23 | 0.0 | [0.0;105.0] | 19 | 40.0 | [0;200.0] | 19 | 0.0 | [0.0;67.5] | 0.556 |
| Walking physical activity (mins/week) | 20 | 420.0 | [257.5;577.5] | 16 | 420.0 | [180.0;840.0] | 15 | 18.0 | 631.4 | 0.914 |
| Sitting (mins/weekday) | 18 | 438.1 | 162.6 | 18 | 371.7 | 165.6 | 16 | -59.7 | 123.0 | 0.071 |
| Total MET-minutes | 19 | 1386.0 | [1100.0;3912.0] | 16 | 2439.0 | [1134.1;5548.5] | 14 | 419.1 | 2584.7 | 0.555 |

*Data presented as mean or median where appropriate with corresponding standard deviation (SD) or interquartile range (IQR). Abbreviations: IQR: inter-quartile range, PP: per protocol, SD: standard deviation.*

**Supplementary Table 19: PP: Changes in clinical and patient-reported outcomes from baseline to 6-month follow-up**

| **Clinical and patient-reported outcomes, PP, N = 23** | **Change from baseline** | | | |
| --- | --- | --- | --- | --- |
|  | **N** | **Mean / Median** | **SD / IQR** | **p-value** |
| Percentage weight change (%) | 23 | -5.81 | 7.25 | 0.001 |
| Weight (kg) | 23 | -5.55 | 7.16 | 0.001 |
| Body mass index (kg/m^2^) | 23 | -1.85 | 2.38 | 0.001 |
| Waist circumference (cm) | 23 | -2.70 | 8.34 | 0.135 |
| Waist-to-height ratio | 23 | -0.02 | 0.05 | 0.128 |
| Fat mass (kg) | 23 | -3.54 | 4.75 | 0.002 |
| Body fat percentage (%) | 23 | -1.72 | 2.62 | 0.005 |
| Visceral fat rating | 23 | -1.0 | [-1.50;0.0] | 0.005 |
| Fat-free mass (kg) | 23 | -1.0 | [-3.20;0.05] | 0.005 |
| Muscle mass (kg) | 23 | -1.90 | 2.75 | 0.003 |
| Bone mass (kg) | 23 | -0.1 | [-0.15;0.0] | 0.005 |
| Total body water (kg) | 23 | -1.0 | [-2.40;-0.05] | <0.001 |
| Total body water percentage (%) | 23 | 1.22 | 2.00 | 0.008 |
| Estimated BMR | 23 | -42.0 | [-113.50;0.50] | 0.003 |
| Systolic blood pressure (mmHg) | 23 | -5.67 | 14.14 | 0.067 |
| Diastolic blood pressure (mmHg) | 23 | 0.0 | [-4.75;3.75] | 0.835 |
| Resting heart rate (bpm) | 23 | -6.48 | 11.11 | 0.011 |
| Bilirubin (umol/L) | 23 | 0.78 | 2.95 | 0.217 |
| Alkaline phosphatase (IU/L) | 20 | -0.70 | 7.15 | 0.667 |
| Alanine transaminase (IU/L) | 18 | -2.0 | [-3.75;1.00] | 0.275 |
| Albumin (g/L) | 23 | 0.91 | 1.73 | 0.019 |
| HbA1C (%) | 22 | -0.10 | 0.23 | 0.057 |
| Fasting glucose (mmol/L) | 22 | -0.15 | [-0.38;0.20] | 0.477 |
| Total cholesterol (mmol/L) | 22 | 0.11 | 0.61 | 0.390 |
| Total cholesterol:HDL ratio | 22 | -0.13 | 0.44 | 0.192 |
| HDL-C (mmol/L) | 22 | 0.03 | 0.26 | 0.568 |
| LDL-C (mmol/L) | 22 | 0.13 | 0.48 | 0.227 |
| Non-HDL-C (mmol/L) | 22 | 0.04 | 0.55 | 0.760 |
| Triglycerides (mmol/L) | 22 | -0.15 | [-0.37;0.08] | 0.065 |
| C-reactive protein (mg/L) | 22 | -0.35 | [-1.45;0.07] | 0.008 |
| PoF - Food available | 18 | -0.8 | [-1.33;-0.50] | <0.001 |
| PoF - Food present | 18 | -1.19 | 1.16 | <0.001 |
| PoF - Food tasted | 18 | -0.86 | 0.91 | 0.001 |
| PoF - Total | 18 | -0.8 | [-1.28;-0.38] | <0.001 |
| CoEQ - Craving control | 18 | 25.12 | 20.60 | <0.001 |
| CoEQ - Craving for sweet | 18 | -13.59 | 12.19 | 0.000 |
| CoEQ - Craving for savoury | 18 | -21.75 | 17.43 | <0.001 |
| CoEQ - Positive Mood | 18 | 12.08 | 18.19 | 0.012 |
| CoEQ - How difficult has it been to resist eating this food during the last 7 days? | 18 | -28.50 | 30.58 | 0.001 |
| IWQoL Lite - Physical function | 19 | 15.43 | 16.52 | 0.001 |
| IWQoL Lite - Self esteem | 18 | 22.02 | 20.69 | 0.000 |
| IWQoL Lite - Sexual life | 19 | 14.80 | 20.86 | 0.006 |
| IWQoL Lite - Public distress | 19 | 0.0 | [0.0;17.50] | 0.037 |
| IWQoL Lite - Work | 19 | 12.83 | 10.92 | <0.001 |
| IWQoL Lite - Total score | 18 | 14.92 | 11.48 | <0.001 |
| EQ-5D-3L Health Today | 19 | 8.32 | 13.54 | 0.015 |
| WEMWBS | 91 | 8.05 | 8.24 | <0.001 |
| PHQ-9 | 18 | -3.67 | 4.79 | 0.005 |
| GAD-7 | 81 | -2.00 | 4.47 | 0.075 |
| PSQI | 21 | -1.95 | 2.13 | <0.001 |
| 6-minute walk test distance (m) | 23 | 45.0 | [17.0;50.0] | 0.007 |
| 6-minute walk test post-test heart rate (bpm) | 23 | -3.00 | 25.35 | 0.576 |
| 6-minute walk test RPE | 23 | 0.0 | [-1.0;0.0] | 0.239 |
| Sit to stand duration (s) | 23 | -0.60 | 1.77 | 0.116 |
| Right hand max (kg) | 23 | 2.81 | 4.26 | 0.004 |
| Left hand max (kg) | 23 | 1.44 | 3.69 | 0.074 |
| Dominant hand max (kg) | 23 | 2.81 | 4.26 | 0.004 |

*Data presented as mean or median where appropriate with corresponding standard deviation (SD) or interquartile range (IQR). Abbreviations: BMR: basal metabolic rate, bpm: beats per minute, CoEQ: Control of Eating Questionnaire, EQ-5D-3L: EuroQol five dimensions three levels, GAD-7: Generalised Anxiety Disorder-7, IQR: inter-quartile range, IWQoL-Lite: Impact of Weight on Quality of Life-Lite, PHQ-9: Patient Health Questionnaire-9, PoF: Power of Food Scale, PSQI: Pittsburgh Sleep Quality Index, PP: per protocol, RPE: Rating of Perceived Exertion, SD: standard deviation, WEMWBS: Warwick Edinburgh Mental Wellbeing Scale.*
